# Meta-analytic findings on reading in children with cochlear implants

**DOI:** 10.1101/2021.03.02.21252684

**Authors:** Yingying Wang, Fatima Sibaii, Kejin Lee, Makayla J. Gill, Jonathan L. Hatch

## Abstract

This meta-analysis study aims to quantify the group differences in reading skills between children with cochlear implants and their hearing peers and between children with cochlear implants and children with hearing aids (aged between 3 to 18 years old). Of the 5,642 articles screened, 47 articles met predetermined inclusion criteria (published between 2002 and 2019). The robust variance estimation based meta-analysis models were used to synthesize all the effect sizes. Children with cochlear implants scored significantly lower than their hearing peers in phonological awareness (g = - 1.62, p < .001), vocabulary (g = -1.50, p < .001), decoding (g = -1.24, p < .001), and reading comprehension (g = -1.39, p < .001), but not for fluency (g = -.67, p = .054). Compared to children with hearing aids, children with cochlear implants scored significantly lower in phonological awareness (g = -.30, p = .028). The percentage of unilateral cochlear implant negatively impacts the group difference between children with cochlear implants and their hearing peers. Findings from this study confirm a positive shift in reading outcomes for profoundly deaf children due to cochlear implantation. Some children with cochlear implants may need additional supports in educational settings.

---

Learning to read plays an essential role in a child’s social and cognitive development, and children who read well are more likely to achieve educational and vocational success (Butler et al., 1985; Kern & Friedman, 2009). Many children who are deaf and hard of hearing (DHH) face serious challenges in accessing the auditory information of reading (i.e., phonological processing and letter-sound correspondences) when learning to read (Herman, Roy, & Kyle, 2017). Since the early 1990s, the expansion of Early Hearing Detection and Intervention (EHDI) programs has dramatically increased the number of infants identified with hearing loss within the first year of life in the United States (CDC, 2017). As a result, more infants with hearing loss have received early diagnosis and treatment such as cochlear implantation and hearing-aid fitting. A review article reported that most children who are DHH successfully develop spoken language after cochlear implantation (Ganek, McConkey Robbins, & Niparko, 2012). Many studies reported that children with cochlear implants (CIs) had improved speech perception and production skills (Blamey et al., 2001; A. Geers et al., 2008; Niparko, Tobey, Thal, Eisenberg, Wang, Quittner, Fink, et al., 2010; Wang et al., 2018; Yoshinaga-Itano et al., 2018). Given the positive relationship between spoken language and reading development in children with typical hearing (TH) (Snowling & Melby-Lervåg, 2016), it is reasonable to suggest that DHH children may benefit from using cochlear implants when learning to read. However, there is considerable variability in cochlear implantation benefits on various emergent and later reading skills.

Learning to read occurs long before children begin to receive formal reading instruction in school. During early childhood, phonological awareness (PA) is critical for reading development in children with TH and has been suggested to be a strong predictor of reading outcomes (Shanahan & Lonigan, 2010). PA is an umbrella term representing the ability to abstract and manipulate segments of spoken language (i.e., blending or deletion) (Spencer & Tomblin, 2009). Despite the early access to spoken language through cochlear implantation, children with CIs still scored significantly lower than children with TH on PA tasks (Bell et al., 2019; Johnson & Goswami, 2010; Nittrouer, Lowenstein, & Holloman, 2016b). Compared to children with hearing aids (HAs), children with CIs had significantly lower scores for the rhyme task, but equivalent scores in the syllable and phoneme tasks (James, Brown, & Brinton, 2005). However, Nittrouer et al. (2012b) found both groups of children with hearing loss had significantly lower scores on phoneme tasks than their hearing peers and no significant differences in PA skills between 25 children with CIs and 8 children with HAs. The lack of differences in PA skills between the CI and HA groups might be due to the small sample size of the HA group.

The developmental sequence of learning to read for children with TH holds for children who are DHH, based on the report from National Center for Special Education Research (Connor et al., 2014) and the Qualitative Similarity Hypothesis (QSH) (Paul & Alqraini, 2019). For instance, the Simple View of Reading (SVR), as an influential model for children with TH, supports that reading comprehension is not a serial process with decoding preceding linguistic comprehension but rather interactive (“product”) processes that involve both decoding and linguistic comprehension (Gough & Tunmer, 1986). The SVR also holds for children who are DHH (Beverly Trezek & Mayer, 2019). Vocabulary knowledge, viewed as part of oral language, is a key linguistic skill that supports reading development in both children with TH (Muter et al., 2004) and children who are DHH (Easterbrooks et al., 2008; Geers, 2003; Worsfold et al., 2018). Children with TH develop vocabulary receptively and expressively. Receptive language vocabulary refers to words understood when they are heard or read (Vatalaro et al., 2018). Expressive language vocabulary refers to words expressed aloud. As early as 12 months, typically developing infants are learning new words rapidly and have 200-300-word expressive vocabulary on average by the age of three (Fenson et al., 1994). Children with CIs usually gain access to auditory inputs and learn spoken language at least a year later than their hearing peers. Some studies found children with CIs scored significantly lower than children with TH in receptive vocabulary tasks (Ambrose, 2009; Bell et al., 2019), while Nittrouer et al. (2018) demonstrated that children with TH were outperformed children with CIs in expressive vocabulary tasks. Moreover, Schorr et al. (2008) identified significantly lower performance in both receptive and expressive vocabulary tasks for children with CIs when compared to their hearing peers. A recent meta-analysis evaluated 12 articles and also supported that children with CIs scored significantly lower than their hearing peers in both vocabulary tasks (Lund, 2016). On the contrary, other studies have reported that children with CIs did not perform significantly differently in receptive and expressive vocabulary tasks than their hearing peers (Luckhurst et al., 2013; Wechsler-Kashi et al., 2014). The contradicting findings may be due to the broad age range in these study samples, which led to more significant variabilities that washed out the group differences.

Based on the SVR, the decoding skill is foundational for reading to learn. Children with TH who master the decoding skill are not necessarily good at reading comprehension. In addition, those children with TH who are poor decoders often perform poorly in reading comprehension. Nevertheless, the early decoding skill predicated later reading comprehension in children with TH (Foorman et al., 2018; Kendeou et al., 2009). Similarly, the decoding skill was also a good predictor for later reading skills in children with CIs (Bell et al., 2019; Mayer, 2007). The decoding skill refers to the process of identifying real words and/or pseudowords timed or untimed (Foorman et al., 2018). If children become more automatic in decoding, they can pay more attention to comprehend what they read. Weisi et al. (2013) found 24 second-grade Persian-speaking children with CIs scored significantly lower on word reading tasks than their hearing peers and failed to gain similar improvement in word reading as their hearing peers in a year. Similarly, Henricson et al. (2012) 33 children with CIs (mean age of 9 years old) scored significantly lower than both 43 children with HAs and 120 children with TH on a word-spotting task during which participants were asked to push a button as soon as they identified a real word among series of nonwords. On the contrary, Nakeva von Mentzer et al. (2013) reported 17 younger children with CIs (5-7 years old) performed equally to 16 of their hearing peers but scored significantly higher in the letter-naming task than 15 children with HAs. The mixed findings can be due to the variability in hearing loss characteristics among children with HAs and the study sample’s age range.

Fluency represents the rate, accuracy, and prosody of reading and is identified as a critical component of reading. Fluency has been positively correlated with reading comprehension and is a part of the developmental process of building decoding skill that forms a bridge to reading comprehension in typically developing children (Pikulski & Chard, 2005; Therrien, 2004). Historically, fluency was a neglected aspect of reading until the early 2000s (National Institute of Child Health and Human Development, 2000). Thus, little research on fluency in children who are DHH is available. The rapid automatized naming (RAN) task provides insight into fluency and predicts later growth in fluency (Lervåg & Hulme, 2009; Norton & Wolf, 2012). In this meta-analysis, the RAN task was included as a fluency measure to include more studies possibly. Children with CIs (mean age: 6.8 years) performed equally well in RAN compared to children with TH (Lee et al., 2012; Nittrouer et al., 2012b), whereas children with CIs (age range from 5 to 14 years) performed below their hearing peers in RAN (Bell et al., 2019; Schorr et al., 2008; Soleymani et al., 2016; Weisi et al., 2013). It is unclear to what extent children with CIs differ from their hearing peers and children with HAs in fluency construct.

The ultimate goal of reading for children is to comprehend the written text and successfully transition from learning-to-read to reading-to-learn. If children with CIs lag in foundational skills compared to children with TH or children with HAs, children with CIs are at risk for slow decoding of single words and slow vocabulary learning. Consequently, children with CIs are expected to perform below their hearing peers in reading comprehension tasks. Vermeulen et al. (2007) reported that 50 Dutch children with CIs (mean age of 12.8 years) performed reading comprehension tasks significantly better than 504 deaf Dutch children without CIs and more poorly than 1,475 Dutch children with TH (mean age of 10.1 years) over four grade levels. On average, children with CIs scored -3.6 standard deviation below the norm, which is much better than deaf children without CIs (-7.2 standard deviation below the norm). In addition, 25-50% of children with CIs at each grade level scored within or above the 95% confidence interval of the children with TH. However, a recent review paper evaluated 21 studies published between 1996-2016 regarding reading comprehension in 1000 children with CIs and summarized that most children with CIs scored within the average range in standardized reading comprehension tests (Mayer & Trezek, 2018). They also indicated a wide range of variability in performance among children with CIs, but the review paper did not quantitatively synthesize the 21 studies’ results.

The current meta-analysis focuses on the above mentioned five constructs, including PA, vocabulary (receptive and expressive), decoding, fluency, and reading comprehension, which were identified as five critical components of reading in children with TH by the report of the National Reading Panel (National Institute of Child Health and Human Development, 2000) and have guided other research (Nation & Snowling, 2004; Petrill et al., 2006; Pikulski & Chard, 2005; Silverman et al., 2013; Suggate, 2016; Therrien, 2004). Other factors such as background knowledge, inferencing, executive function, and motivation are also important for reading development, but are beyond this meta-analysis scope.

In summary, previous studies on how children with CIs performed in emergent and later reading skills compared to children with TH and children with HAs have produced mixed findings. QSH suggested that the reading development for children who are DHH is “qualitatively similar but quantitatively delayed” compared to their hearing peers (Paul, 1998; Trezek et al., 2010). However, it remains unclear to what extent children with CIs differ from children with TH and children with HAs in emergent and later reading skills. The current meta-analysis aims to quantitatively synthesize studies’ results by conducting secondary statistical analyses on reading skills between children with CIs and children with TH, and between children with CIs and children with HAs. Meta-analysis effectively pools the studies’ results to increase statistical power applied to research questions (Hedges et al., 2010).

The conflicting findings and the wide range of score variability in children with CIs can be attributed to considerable differences in participants’ age at testing, age at onset of deafness, age at cochlear implantation, duration of implant use, and percentage of unilateral CI users. The five factors may be possible moderators affecting the group differences in reading skills between children with CIs and their hearing peers/children with HAs.

Children with CIs face more challenges in reading over time than their hearing peers (Geers et al., 2008; Sarant et al., 2015). Geers et al. (2008) reported that the reading comprehension gap between children with CIs and their hearing peers increased with age. Van der Kant et al. (2010) also demonstrated that the deviation of a reading comprehension measure from the norm in Flemish and Dutch children with CIs increased with age. In addition, Nittrouer et al. (2018) reported that group differences in PA, vocabulary, and decoding between the CI and HA groups remained somewhat similar across the second and sixth grades. Thus, the gap in reading skills between children with CIs and their hearing peers may widen with age.

Research has shown that children whose deafness occurred postlingually acquired higher speech perception ability than children with prelingual deafness (Ahmad et al., 2012; Buckley & Tobey, 2011; Busby et al., 1993; Hinderink et al., 1995; Ruff et al., 2017; Seldran et al., 2011). Moreover, deaf children who had normal hearing for even a short period after birth and received a CI shortly after losing their hearing had better speech and language proficiency (Geers, 2004; Niparko & Geers, 2004). Thus, the age at onset of deafness may negatively affect the group difference in reading skills.

The age at implantation is usually negatively correlated with reading skills. Earlier implantation was associated with higher reading skills in children with CIs, and the reading age of children implanted before 42 months matched their chronological age (Archbold et al., 2008). The early implanted CI group performed better on vocabulary measures than the later implanted CI group (Johnson & Goswami, 2010). However, Harris et al. (2011) did not find significant decoding differences between an early-implanted group and a later-implanted group. The advantage of earlier implantation might stem from a shorter period of auditory deprivation. More extended auditory deprivation could lead to cross-modal neural plasticity, causing the primary auditory cortex to be used by other sensory inputs (Lazard et al., 2012; Sharma et al., 2009). If a CI is received later than 42 months, the benefits are limited (Lazard et al., 2012). Before 42 months, neural plasticity is optimal, and thus a normal range of latency of the first positive peak of the cortical auditory evoked potential post-CI is achievable (Sharma et al., 2002).

The duration of implant use has also been studied as a possible factor affecting speech recognition outcomes (Dillon et al., 2012; Geers, 2004; Nittrouer et al., 2012b). Studies have identified a positive correlation between implant use duration and phonemic awareness in children with CIs (Dillon et al., 2012; Nittrouer et al., 2012b).

Studies have shown that children who received bilateral CIs performed better in language comprehension, vocabulary, and reading comprehension than children who received a unilateral CI (Boons et al., 2012; Ching et al., 2014; Sarant et al., 2015). Bilateral CIs are superior in sound localization and spatial release masking to a unilateral CI, which should enhance learning the language by helping children hear speech in the environment more efficiently (Boons et al., 2012; Sarant et al., 2015). However, some studies did not find that bilateral CIs significantly benefit language development (Niparko et al., 2010; Nittrouer et al., 2012b). The signal processing is the same for the bilateral CIs, and the acoustic structure that underlies phonemic categories is similarly represented to the brain through both CI devices (Nittrouer et al., 2012b). This statement is valid under the circumstance that symmetrical insertion depth between two ears is guaranteed, and there are no frequency-place mismatches between ears through CI device programming. Otherwise, the spectral content that is perceived can be different between bilateral CIs and unilateral CI. In addition, the differences in neural health patterns between ears before implantation can affect phonemic awareness after implantation. If children had a unilateral CI in the ear with poorer neural health and more spectral smearing, the phonemic categories’ acoustic structure would not be represented. Thus, the prediction could be made that the higher percentage of children with a unilateral CI will increase the group differences in reading skills between children with CIs and their hearing peers.

The main objective of the current meta-analysis is to quantitatively summarize the group differences in five constructs (PA, vocabulary, decoding, fluency, and reading comprehension) between children with CIs and their hearing peers and between children with CIs and children with HAs. The mixed findings in the current literature need to be further analyzed to answer the following questions: (1) do reading skills (a composite of all five constructs) significantly differ between children with CIs as compared to children with TH? (2) do reading skills (a composite of all five constructs) significantly differ between children with CIs as compared to children with HAs? (3) do the group differences between children with CIs as compared to children with TH vary among constructs? (4) do the group differences between children with CIs as compared to children with HAs vary among constructs? (5) does vocabulary domain (receptive versus expressive) alter the magnitude of group differences between children with CIs as compared to children with TH? (6) Does age at testing, age at onset of deafness, age at implantation, duration of implant use, or percentage of unilateral CI users significantly relate to the magnitude of group differences between children with CIs as compared to children with TH?

The present study will quantify the group differences in reading skills between children with CIs and their hearing peers, and between children with CIs and HAs. Our findings will highlight the scale of reading challenges that children with CIs face. Children with CIs who are a new generation of children who are DHH benefit from early identification, early treatment, early intervention programs (i.e., speech therapy, parent education), and advances in cochlear implant devices. They attend mainstream education, and their special need for support may be overlooked. Our findings may give educators a better expectation on reading skills for children with CIs and subsequently provide appropriate interventions to prevent them from falling behind their hearing peers.

## Material and Methods

### Search Strategy

A Preferred Reporting of Systematic Reviews and Meta-Analyses (PRISMA) flow diagram (Moher et al., 2009) outlined a comprehensive online search. We searched Academic Search Premier, PsycArticles, PsycInfo, ERIC, PubMed, Open Dissertations, ProQuest Dissertations, and Theses Global (Figure 1). Open Dissertations, ProQuest Dissertations, and Theses Global were used to search for unpublished work for minimizing potential publication bias. The titles and abstracts were searched using the following key terms: “hearing loss” OR “cochlear implant” OR “hearing aid” OR “deaf” OR “hard of hearing” and “reading” OR “phonological” OR “phonology” OR “phonemic” OR “word identification” OR “word attack” OR “naming” OR “RAN” OR “vocabulary.” Publications after 1990 were included because CIs were not commercially available for children until 1990 in the United States.

**Figure 1.**
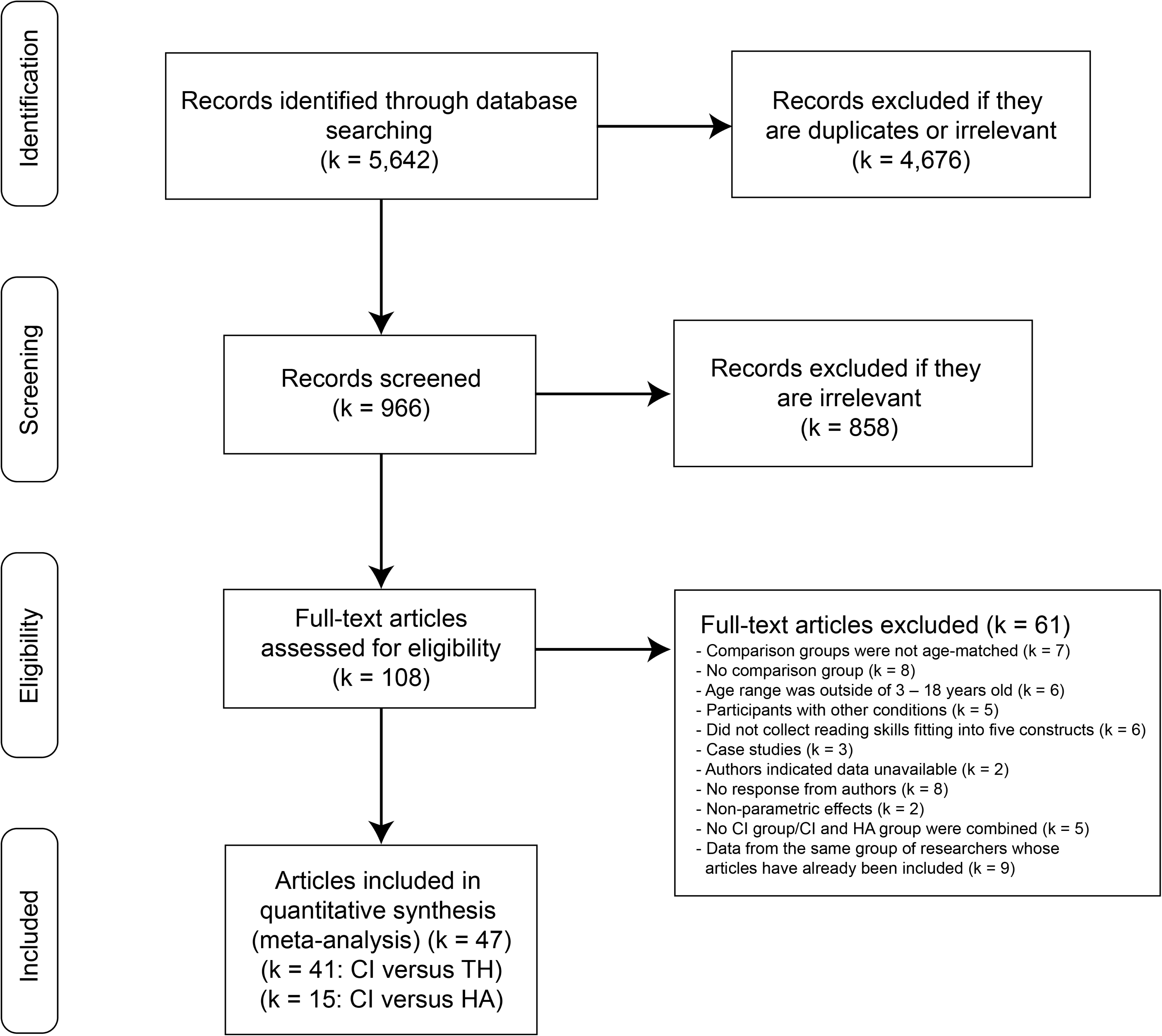
Preferred Reporting of Systematic Reviews and Meta-Analyses. k is the number of articles.

The initial search yielded 5,642 articles, and 966 unique abstracts were included for further screening after removing duplications. 858 irrelevant articles were removed, and there were 108 articles left to be further assessed for eligibility.

### Study Selection

The inclusion criteria included: (1) age range from 3 to 18 years old, (2) comparison between individuals with CIs and those with TH, or comparison between individuals with CIs and those with HAs, (3) measures of reading skills such as phonological processing, word recognition, vocabulary, reading comprehension, fluency, etc. The exclusion criteria were: (1) individuals with no additional disabilities (i.e., Autism Spectrum Disorder, Attention Deficit Hyperactivity Disorder, visual impairment, and cognitive disabilities, etc.) (2) case studies, (3) review articles, (4) written in any languages other than English (not excluded if they were conducted in a language other than English (i.e., 21 of the 41 articles comparing children with CIs to their hearing peers and six of the 15 publications comparing children with CIs to children with HAs were carried out in languages other than English). Some articles that did not include a typical-hearing comparison group or only compared children with CIs to published test norms were excluded. Missing a comparison group or using test norms does not allow researchers to control other confounders other than chronological age. Meta-analysis uses effect sizes to compare group differences, and effect size cannot be effectively computed without a comparison group. Thus, a comparison group was essential for article inclusion to determine to what extent children with CIs differ from their hearing peers and children with HAs using meta-analysis techniques. Thirteen authors were contacted if articles indicated that needed data were collected but not reported. Three authors responded. For the comparison between CI and TH groups, 41 articles were included. Seven articles reported data from children across different grades. Seven articles were from the same research group and used the same cohorts of children. There were 43 independent samples from 41 articles. For the comparison between CI and HA groups, 15 articles and 19 independent samples were included. There were 9 studies included in both CI versus TH and CI versus HA, resulting in 47 articles included in the present study (Figure 1). All 47 articles are listed in Supplementary Table A.2, and all 61 articles excluded due to various reasons are listed in Supplementary Table A.3. In Supplementary Table A.2, the studies conducted in the U.S. reported that most students were in mainstream schools. Most studies indicated their children with CI or HA used oral or total communication mode.

### Data Extraction

The data extracted from each study included participant demographics and psychometric measures of reading skills. Additional data about children who are DHH were extracted, including the age when hearing loss was identified, level of unaided hearing thresholds, age when CIs were activated or implanted, age when HAs were fitted, duration of CIs/HAs usage, and the percent of participants utilizing a unilateral CI. For reading skills, we extracted the tasks’ names and standardized scores or normalized scores of assessments, and raw scores or percent correct scores if standardized scores were not provided. The reading skills were further categorized into five constructs (Supplementary Table A.2): (1) PA (sound manipulation, i.e., rhyming, elision, first-sound matching, etc.), (2) decoding (visual word recognition, irregular word reading, regular word reading, nonword recognition, etc.), (3) vocabulary (expressive and receptive vocabulary), (4) fluency (reading rate subtest, fluency qualitative reading inventory, and rapid automized naming), (5) reading comprehension (sentence completion, passage comprehension, etc.). The second author coded all studies, and the fourth author independently coded 80% of the studies. Then, accuracy was calculated by comparing all quantitative information extracted from studies, including moderators and demographics. The overall accuracy of coding demographic information for studies comparing children with CIs with their hearing peers was 94%. The overall accuracy of coding demographic information for studies comparing children with CIs to children with HAs was 96%. The accuracy of the reading skills for studies comparing CI and TH groups was 96%. The accuracy of the reading skills for studies comparing CI and HA groups was 98%. Any discrepancies in the coding were resolved by re-evaluating the full text and discussions between the two coders.

### Effect Size Computation

Hedges’ g with bias correction (Hedges, 1981) was used to examine the mean differences in reading skills between groups (CI vs. TH, and CI vs. HA) because it was recommended for smaller sample sizes (N < 50), and it provided an unbiased and conservative measure. For studies that reported the group differences, we used the reported means and standard deviations to compute the effect-size estimates expressed as Hedges’ g. For studies that did not report the group differences directly, t-test statistics between groups were used to compute Hedges’ g (Wilson, 2014). The magnitude of standardized mean difference estimates was interpreted based on the following scale (Cohen, 1992): < .20 = no effect, .20 - .49 = small effect, .50 – 0.79 = moderate effect, ≥ 80 = large effect. Within the study, there were multiple psychometric measures of reading skills that were not from independent samples. Therefore, we used robust variance estimation (RVE) to synthesize Hedges’ g without averaging across effect sizes or selecting only one effect size per study (Hedges et al., 2010). Because there were different constructs within the same study sample, we used the correlation weight with moderate ρ = .40 to combine effect-size estimates and model the random effects of between-study variance with consideration of correlated constructs from the same study sample (Hedges et al., 2010). The robumeta R package (Fisher et al., 2017) was used for a random-effects model with RVE to examine the magnitude of standardized mean difference estimates. Both *I*^2^ and *τ*^2^ were calculated to assess the degree of between-study heterogeneity.

### Moderator Analyses

Seven moderators were examined for group comparison between the CI and TH groups, including (1) constructs of reading skills, (2) subtypes of vocabulary, (3) CI group’s age at testing (TH group had matched age at testing), (4) CI group’s age at onset of deafness, (5) CI group’s age at implantation, (6) CI group’s duration of implant use, (7) CI group’s percent of children with unilateral CI. Only one moderator, constructs of reading skills, was examined for group comparison between the CI and HA groups due to the limited number of studies. For the construct moderator, we conducted RVE meta-regression with a small-sample adjusted F-test (Tipton & Pustejovsky, 2015) to evaluate the potential influence of different moderators on the mean group difference in reading skills using the robumeta (Fisher et al., 2017) and clubSandwich R packages (Pustejovsky, 2017). There were five constructs of reading skills, and we were interested in testing whether the magnitude of standardized mean difference estimates depended on the different constructs of reading skills. Because we were interested in the effect of each construct, we then built random-effects RVE models for each construct separately. There were two subtypes of vocabulary, including receptive and expressive vocabulary. The current meta-analysis also examined whether the group differences between the CI and TH groups depended on the different subtypes of vocabulary (receptive and expressive). The statistical significance level was set at p < .05.

### Publication Bias and Sensitivity Analysis

We searched for unpublished work to reduce potential publication bias. Publication bias is due to the fact that studies with relatively large effect sizes are more likely to be published than studies with smaller effect sizes. In the current meta-analysis, we aggregated effect sizes per each independent sample using the R package MAd (del Re & Hoyt, 2014). We also generated forest plots and funnel plots of the aggregated effects for visual inspection. To test whether the funnel plot is asymmetric, we used Egger’s test (Egger et al., 1997) of standardized mean differences. In addition, if the funnel plot was asymmetric, Rosenthal’s Fail-safe N was computed to check the number of studies needed to nullify the effect (Rosenthal, 1979).

### Outlier and Sensitivity Analysis

We examined potential outliers using an influential study analysis to determine any influential outliers (Viechtbauer & Cheung, 2010). Additionally, we checked whether the results were affected by different correlation coefficients (e.g., correlation coefficient other than .40) between pairs of effect sizes using the robumeta R package (Fisher et al., 2017).

## Results

### CI versus TH

#### Search results and study characteristics

A total of 41 articles with 43 independent samples and 134 effect sizes were included in comparing CI and TH groups (Supplementary Table A.4 and Supplementary Table A.1). The total number of unique participants in the CI group was 911, and the total number of unique participants in the TH group was 2,438. Supplementary Table A.4 summarizes demographic information, and means were computed from all reported studies.

#### Group Differences in Reading Skills

With the RVE random-effects model, the significant effect of the group difference in reading skills between individuals with CIs and their hearing peers was -1.14 (p < .001, 43 independent samples and 134 effect sizes, 95% confidence interval [-1.45 -.83]), indicating that the CI group scored statistically significantly lower on reading skills than their TH peers. There were substantial levels of between-study heterogeneity (*τ*^2^ = .74, *I*^2^ = 89%). Supplementary Table A.5 summarizes the descriptive statistics of each construct. In addition, table 1 summarizes the univariate RVE models for each construct. The significant effect of the group difference in PA between children with CIs and their TH peers was -1.62 (p < .001), indicating that the CI group scored statistically significantly lower in PA tests than their TH group. The large effect of the group difference in vocabulary between CI and TH was -1.50 (p < .001), indicating that the CI group scored statistically significantly lower on vocabulary tests than their TH group. Furthermore, only expressive vocabulary knowledge showed significant group differences between the CI and TH group (g = -1.57, p < .001). The group differences in reading comprehension between individuals with CIs and their TH peers was -1.39 (p < .001), indicating that the CI group scored statistically significantly lower in reading comprehension tests than their TH peers. The group differences in decoding between CI and TH groups were -1.24 (p < .001). The group differences in fluency were not significant, and the CI group scored marginally lower in fluency tasks than their TH peers (g = - .67, p < .001).

**Table 1.**
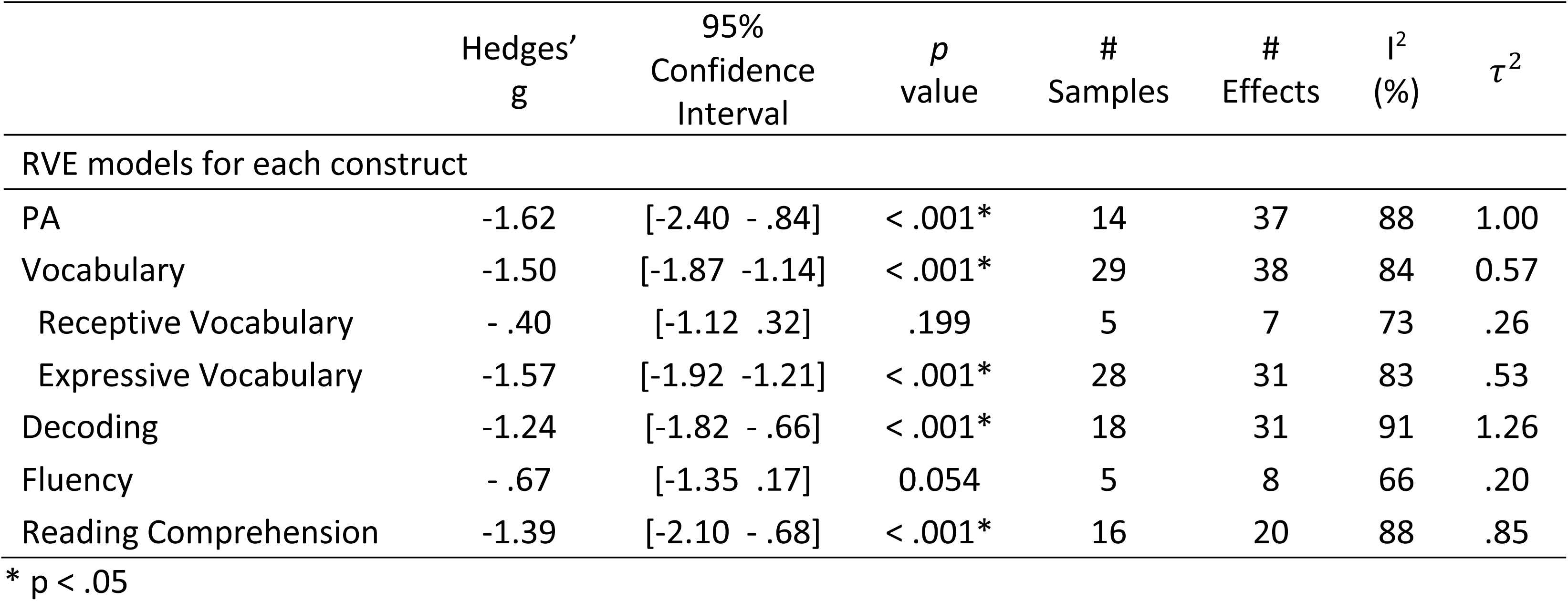
RVE models of each construct for CI versus TH

#### Moderators

Table 2 summarizes the moderator analyses. The construct of reading skills was not a significant moderator for the group differences between CI and TH (F = 1.76, p = .192), suggesting that the magnitude of standardized mean difference estimates between CI and TH groups were not affected by the type of constructs of reading skills. The vocabulary subtype was a significant moderator for the group differences between CI and TH (F = 15.4, p = .015), indicating that the magnitude of standardized mean difference estimates between CI and TH groups were affected by the subtype of vocabulary knowledge. The CI group’s age at testing, age at onset of deafness, age at implantation, and implant use duration were not significant moderators for the group differences between CI and TH (Table 2). The CI group’s percent of children with unilateral CI was a significant moderator for group differences between CI and TH (p = .008), indicating more children with unilateral CI in the CI group negatively impacted the group differences between CI and TH.

**Table 2.**
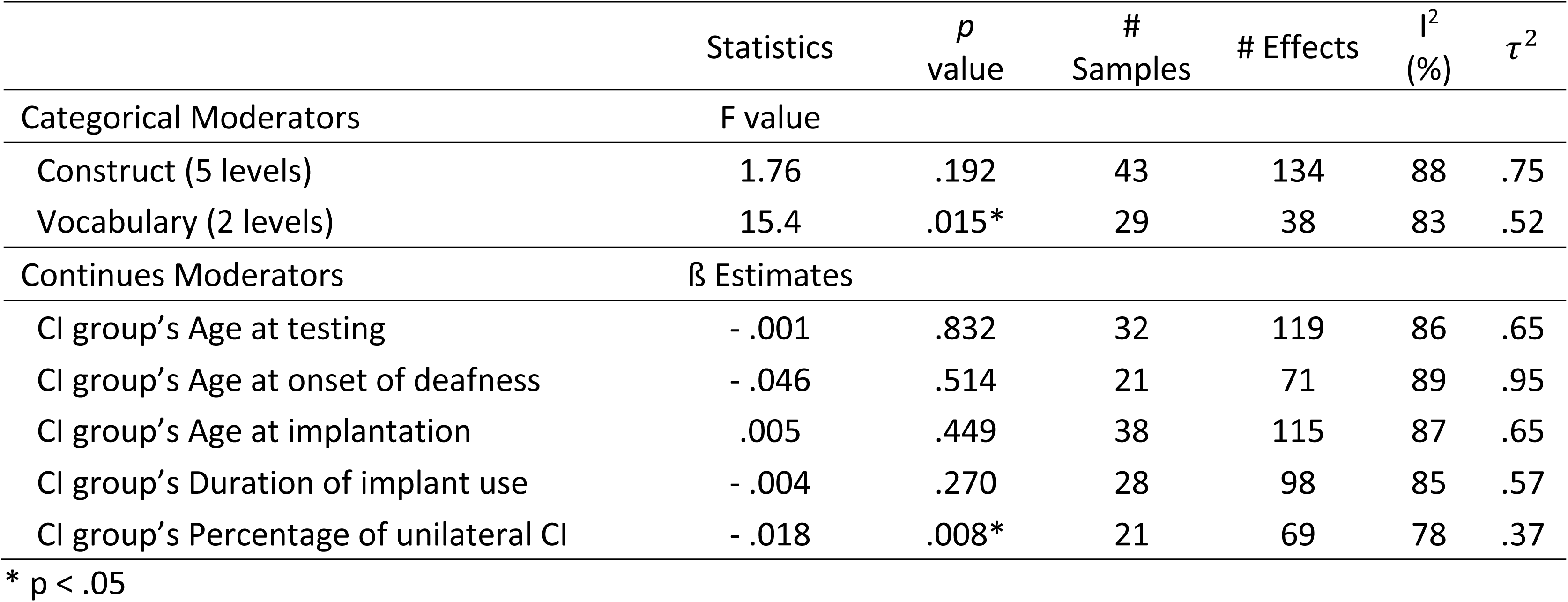
Moderator Analyses for CI versus TH

| | Statistics | *p*<br>value | #<br>Samples | # Effects | *I*<sup>2</sup><br>(%) | $\tau^2$ |
| --- | --- | --- | --- | --- | --- | --- |
| Categorical Moderators | F value |  |  |  |  |  |
| Construct (5 levels) | 1.76 | .192 | 43 | 134 | 88 | .75 |
| Vocabulary (2 levels) | 15.4 | .015* | 29 | 38 | 83 | .52 |
| Continues Moderators | $\beta$ Estimates | | | | | |
| CI group's Age at testing | - .001 | .832 | 32 | 119 | 86 | .65 |
| CI group's Age at onset of deafness | - .046 | .514 | 21 | 71 | 89 | .95 |
| CI group's Age at implantation | .005 | .449 | 38 | 115 | 87 | .65 |
| CI group's Duration of implant use | - .004 | .270 | 28 | 98 | 85 | .57 |
| CI group's Percentage of unilateral CI | - .018 | .008* | 21 | 69 | 78 | .37 |

\* $p < .05$

#### Publication bias

Studies with significant findings, large effects, or large sample sizes were more likely to be published (Hedges, 1989). If the effect size is big, a result is more likely to become statistically significant for any sample size. Particularly for small sample sizes, very large effect sizes are needed to reach statistical significance. We searched the unpublished work and tried to minimize publication bias. We generated both a forest plot (Supplementary Figure A.1) and a funnel plot (Supplementary Figure A.3). The funnel plot indicated a slight asymmetry, with a majority of the smaller studies clustering to the left of the mean. The visual impression is confirmed by Egger’s test, which showed a statistically significant asymmetry in the funnel plot (t = -7.24, p < .001), suggesting the presence of small-study effects. As a whole, the smaller sample-size studies did tend to report a large group difference between children with CIs and their TH peers (see the forest plot in Supplementary Figure A.1). Rosenthal’s Fail-safe N was 76,363, suggesting that there would need to be 76,363 studies added to the current meta-analysis before the cumulative effect would become statistically non-significant. Given that this meta-analysis identified only 41 articles that met the inclusion criteria, it is unlikely that nearly 76,363 studies were missed. While we may have overstated the group differences in reading skills between children with CIs and their TH peers, it is unlikely that the actual group difference is zero.

#### Outliers and sensitivity

There were three influential cases (Rezaei et al., 2016; Soleymani et al., 2016; Weisi et al., 2013). But the removal of these three influential cases did not affect the statistical inference of the large effect of group differences in reading skills (g = -1.08, p < .001, 95% confidence interval [-1.36, - .81]), or the results of moderator analyses and publication bias analysis. Therefore, we included all the effect sizes in the current meta-analysis of group differences in reading skills between CI and TH. In addition, the large effect of the group differences in reading skills between individuals with CIs and their TH peers did not change when degrees of correlations varied from 0 to 1 (e.g., 0, .20, .40, .60, .80, and 1.00).

### CI versus HA

#### Search results and study characteristics

A total of 15 articles with 19 independent samples, and 59 effect sizes were included in the comparison between CI and HA groups (Supplementary Table A.6 and Supplementary Table A.1). The total number of unique participants in the CI group was 448, and the total number of unique participants in the HA group was 434. Supplementary Table A.6 summarizes demographic information and means computed from all reported studies.

#### Group Differences in Reading Skills

With the RVE random-effects model, the small effect of the group differences in reading skills between children with CIs and children with HAs was -.04 (p = .726, 59 effect sizes, 95% confidence interval [-.25 -.17]), indicating that there were no statistically significant differences between CI and HA groups in reading skills. There were moderate levels of between-study heterogeneity (*τ*^2^ = .15, *I*^2^ = 61%). In addition, table 3 summarizes the RVE models for each construct. The group difference in PA between CI and HA was - .30 (p = .028, 13 effect sizes, 95% confidence interval [-.55 -.06]), indicating that the CI group scored statistically significantly lower on PA tests than the HA group. The group differences associated with vocabulary, decoding, fluency, and reading comprehension were - .08 (p = 0.692), - .24 (p = .179), -.24, and .08 (p =.568), respectively. The group differences in vocabulary, decoding, fluency, and reading comprehension between children with CIs and children with HAs were not statistically significant.

**Table 3.** RVE models of each construct for CI versus HA

| | Hedges'<br>g | 95%<br>Confidence<br>Interval | *p*<br>value | #<br>Samples | #<br>Effects | I<sup>2</sup><br>(%) | $\tau^2$ |
| --- | --- | --- | --- | --- | --- | --- | --- |
| RVE models for each construct |  |  |  |  |  |  |  |
| PA | -.30 | [-.55 -.06] | .028* | 6 | 13 | 0 | .00 |
| Vocabulary | -.08 | [-.50 .34] | .692 | 13 | 15 | 79 | .43 |
| Decoding | -.24 | [-.61 .13] | .179 | 11 | 18 | 65 | .22 |
| Fluency<sup>+</sup> | -.24 | N/A | N/A | N/A | N/A | N/A | N/A |
| Reading Comprehension | .08 | [-.22 .38] | .568 | 11 | 11 | 53 | .10 |

\* $p < .05$ , <sup>+</sup> degree of freedom < 4 and RVE result was not reliable.

### Moderator

Due to the small number of studies included in this meta-analysis, only one moderator (constructs of reading skills) was examined. The construct of reading skills was not a statistically significant moderator for the group differences between CI and HA (F = 2.38, p = .127).

### Publication bias

To assess publication bias, we generated a forest plot (see Supplementary Figure A.2) and a funnel plot (see Supplementary Figure A.4). The Egger’s test of asymmetry was not significant (t = -.51 p = .611), indicating no significant publication bias. Since Egger’s test was not significant, Rosenthal’s Fail-safe N was not computed.

### Outlier and Sensitivity Analyses

There was one influential case (Henricson et al., 2012). The influential case did not affect the statistical inference of the group differences in reading skills (g = -.02, p = .806, 95% confidence interval [-.22, .17]) or the results of publication bias analysis and moderator analyses. Therefore, we included all the effect sizes in the current meta-analysis. Additionally, the group difference in reading skills between CI and HA did not change when degrees of correlations varied from 0 to 1.

## Discussion

The current study is the first comprehensive and quantitative meta-analysis to pool effect sizes of reading skills between children with CIs and their hearing peers, and between children with CIs and children with HAs. The current meta-analysis systematically examined to what extent children with CIs differ in reading tests than their hearing peers or children with HAs. Findings from this meta-analysis provide rigorous insights regarding group differences (CI versus TH and CI versus HA) on test performance measuring emergent and later reading skills, which have implications not only for informing educators what to expect for children with CIs when it comes to emergent and later reading skills, but also for better understanding of reading challenges for children who are DHH.

Overall, when five constructs (PA, vocabulary, decoding, fluency, and reading comprehension) were included in the RVE random-effect model (a total of 134 effect sizes included), there was a significant large effect of group differences between children with CIs and their hearing peers. On average, children with CIs scored significantly lower than their hearing peers, but the large effect only indicated children with CIs performed one standard deviation (SD) below their hearing peers on tests measuring their emergent and reading skills. This result aligns with those studies reporting that children with CIs scored significantly lower than their hearing peers, but still achieved within the normal range on reading tests (i.e., out of normal range is defined as more than 1.5 SD or more below the norm) (Dillon et al., 2012; Spencer et al., 2003). While the construct of reading skills did not significantly change the magnitude of the group differences between children with CIs and their hearing peers, our results also indicated children with CIs struggled more in PA, vocabulary, and reading comprehension and less in decoding and fluency. Our findings align with the QSH (Paul & Alqraini, 2019) and suggest that similar to their hearing peers, children with CIs require the same foundational skills to learn to read. The average performance among children with CIs in reading skills in the current meta-analysis is encouraging and confirms a positive shift in reading outcomes for profoundly deaf children as a consequence of cochlear implantation despite the poor scores in all constructs. Although there is variability among constructs of reading skills, overall reading outcomes exceed the historically reported 4th-grade ceiling (Paul & Alqraini, 2019). Notably, some children with CIs performed on reading tests at an age-appropriate level, consistent with a recent review paper on literacy outcomes that suggested an incredible improvement with many students with CIs achieving reading outcomes at the age-appropriate range (Mayer & Trezek, 2018). Our findings highlight the scales of difficulties in each construct of reading skills faced by children with CIs and provide a rationale for early intervention for children with CIs to develop foundational reading skills (Lederberg et al., 2014).

The findings concerning group differences between children with CIs and children with HAs are less robust due to a limited number of articles included in the current meta-analysis (15 articles, 19 independent samples, and 59 effect sizes). For PA construct only, there was a small effect of group difference between children with CIs and children with HAs, in agreement with those studies reporting children with moderately severe and severe hearing loss who used HAs performed better in PA than children with profound hearing loss who received CIs (Fitzpatrick et al., 2012). In addition, PA intervention studies demonstrated that both children with CIs and children with HAs improved phonological skills after the intervention, and PA intervention given to children with hearing loss who wear CIs and/or HAs during their pre-kindergarten school year equipped them to enter kindergarten with PA skills that generally exceeded minimal entry-level expectations for their hearing peers (Sohail & Nabeel, 2019; Werfel et al., 2016). Therefore, there is a strong rationale for providing early intervention to children who are DHH with functional hearing for the development of their foundational reading skills (Lederberg et al., 2014).

Emergent reading skills start before PA when young children are surrounded by environmental print (Neumann et al., 2012). In pre-kindergarten and kindergarten, instruction emphasizes phonemes, oral language, and some letter-to-sound knowledge (Shanahan & Lonigan, 2010). The largest effects of group differences between children with CIs and their hearing peers and between children with CIs and children with HAs were found in the PA construct. The poor performance in PA is a consequence of impoverished and degraded auditory inputs through CIs, aligned with other studies (Ambrose et al., 2012; Rastegarianzadeh et al., 2014). In the current study, the construct of PA consisted of syllable deletion, syllable counting, initial/final consonant discrimination, phoneme deletion, elision, blending, segmenting, and rhyme tasks. The 95% confidence interval for PA was wide and can be accounted for in various PA tasks included. Children with CIs (mean age: 8.33 years old) performed better on the syllable test than the rhyme test and better on the rhyme test than the phoneme test (James et al., 2005). Children with CIs (mean age: 8.58 years old) scored significantly lower on initial and final phoneme decision tasks than their hearing peers, but not on syllable counting task (Nittrouer et al., 2016a). The CIs provide relatively degraded spectral information and cannot fully normalize a child’s auditory experience. Thus, compared to children with TH, children with CIs experience lower phonological sensitivity (Eisenberg et al., 2004; Harlan et al., 2007; Wei et al., 2004), and delayed speech and language development (Geers et al., 2008; Nittrouer & Caldwell-Tarr, 2016c). Our meta-analysis also demonstrated that children with HAs outperformed children with CIs in the PA tasks less than one standard deviation. Children with HAs exhibit mild to severe hearing loss, while children with CIs exhibit severe to profound hearing loss. The two groups usually consist of heterogeneous cohorts with unmatched hearing profiles (i.e., unaided hearing level, unilateral versus bilateral).

The group difference in vocabulary between children with CIs and their hearing peers was second among all five constructs. The current meta-analysis examined 29 independent samples and found that children with CIs scored 1.5 SD below their hearing peers on vocabulary tasks. Moreover, lower performance in PA tasks in children with CIs could affect vocabulary development. Children with CIs took significantly more days to acquire their first 100 words than their hearing peers (Nott et al., 2009). Meanwhile, the lag in vocabulary in children with CIs could also affect the development of phonological representations according to the lexical restructuring model (Metsala & Walley, 1998). The magnitude of group differences varied across studies that were included in the current meta-analysis. However, children with CIs always performed more poorly on vocabulary tasks than their hearing peers regardless of different types of psychometric tests used to measure vocabulary knowledge, which is in agreement with a recent meta-analysis comparing vocabulary knowledge in children with CIs aged 49 to 109 months versus their hearing peers (Lund, 2016). Lund et al. (2016) evaluated 12 articles using forest plots and found that children with CIs performed more poorly than their hearing peers in both receptive (-.46 to -2.00) and expressive vocabulary (-.34 to -1.06) (Lund, 2016). In addition, we found no significant group difference in vocabulary knowledge between children with CIs and children with HAs, which can be due to the heterogeneity of hearing loss profiles between the two groups.

The difference between receptive and expressive vocabulary in the current meta-analysis reached statistical significance, contrary to the recent meta-analysis (Lund, 2016). The present meta-analysis indicated that expressive vocabulary was more delayed for children with CIs than their hearing peers. The current study found that children with CIs scored 1.6 SD below their hearing peers on expressive vocabulary and .4 SD below their hearing peers on receptive vocabulary. However, Lund et al. (2016) demonstrated that children with CIs scored 20.33 points lower on measures of receptive vocabulary and 11.99 points lower on measures of expressive vocabulary than their hearing peers. The discrepancy can be due to the use of raw scores versus norm-referenced or normalized scores of assessments, and the number of different studies included in each study. Lund et al. (2016) mentioned that more studies would have increased the likelihood of a significant difference between receptive and expressive vocabulary knowledge. The current study included more studies on expressive vocabulary, not on receptive vocabulary. Due to the unbalanced number of studies between receptive (7 effect sizes) and expressive (31 effect sizes) vocabulary knowledge in the current study, poorer expressive vocabulary may reflect properties of a test rather than a quantitative difference in vocabulary knowledge.

The group difference in decoding between children with CIs and their hearing peers was slightly smaller than PA and vocabulary constructs. There was no significant difference in decoding between children with CIs and children with HAs. In the current meta-analysis, decoding included some tasks that require reading words covertly and other tasks that require reading words overtly. For reading words covertly, children with CIs could use strategies other than phonological coding strategies to recognize words visually. For reading words overtly, children with CIs had to use phonological coding strategies to produce words overtly. Thus, the alternative strategies in word-level decoding led to more minor group differences between children with CIs and their hearing peers. Phonological coding strategies for word-level decoding are essential for skilled reading (Bell et al., 2019; Geers, 2003; Watson, 2002), but other visual strategies through semantic cues might help to decode.

The current meta-analysis did not find significant group differences in fluency tasks between children with CIs and their hearing peers, and between children with CIs and children with HAs. But these findings are less robust due in part to the fact that there is a lack of literature on fluency in children who are DHH, with only 8 effect sizes for CI versus TH groups and 2 effect sizes for CI versus HAs. In addition, the RAN task was included in the construct of fluency since researchers suggested that RAN is an early indicator that predicts reading fluency (Norton & Wolf, 2012). There were mixed findings in the studies included in the current meta-analysis, which may contribute to the non-significant group differences. Some studies found that children with CIs performed as well as their hearing peers in RAN tasks (Lee et al., 2012; Nittrouer, Caldwell, & Holloman, 2012a), whereas other studies identified significantly lower performance in RAN tasks for children with CIs than their hearing peers (Bell et al., 2019; Schorr et al., 2008). The mixed results could be attributed to different types of RAN tasks. In general, children learned to name colors and objects first, then numbers and letters (Denckla & Rudel, 1974). For example, Bell et al. (2019) required children with CIs to name letters or numbers, whereas Lee et al. (2012) and Nittrouer et al. (2012a) asked children with CIs to name colors and objects.

The current meta-analysis identified that children with CIs performed 1.39 SD below their hearing peers on reading comprehension tasks. There was no significant difference in reading comprehension between children with CIs and children with HAs. Based on the SVR model (Gough & Tunmer, 1986), it is expected for children with CIs to have lower performance on reading comprehension due to poorer decoding and linguistic comprehension (vocabulary) skills. The cloze tasks used to measure reading comprehension require children to know the syntactic structure of language. Since children with CIs lagged behind their hearing peers in grammatical abilities (Nittrouer et al., 2014; Schorr et al., 2008) and morphosyntactic abilities (Nittrouer et al., 2018), children with CIs would make more mistakes due to a possible deficiency in understanding syntactic structure during cloze tasks. Notably, the large effect of group difference in reading comprehension demonstrated that children with CIs scored 1.39 SD below their hearing peers, within the normal range. This finding agrees with the positive shift in reading outcomes described in a recent review article (Mayer & Trezek, 2018).

Further analysis reveals that the magnitude of difference in emergent and later reading skills between children with CIs and their hearing peers does not significantly relate to chronological age at testing, age at onset of deafness, age at implantation, and implant use duration. We hypothesized that group differences in reading skills between children with CIs and their hearing peers would increase. Because the Matthew effect in reading suggested that reading ability gaps increase with age because the poor pre-reading skills can lead to less reading experience that negatively impacts the reading ability (Stanovich, 1986). In addition, previous literature suggested that students with CIs experience a gap in reading that increases as they get older (Geers & Hayes, 2011; van der Kant et al., 2010). However, the current meta-analysis did not find a significant effect of chronological age at testing on group differences, which can be due to the use of mean age and norm-referenced or normalized scores of assessments in most studies included in the present meta-analysis. The mean age of the CI group was used to represent the whole comparison groups’ mean age because studies used the age-matched controls. Some studies had a small age range within each group (Ambrose, 2009; Nittrouer, 2016a; Rezaei et al., 2016; Weisi et al., 2013), whereas other studies included a wide age range within each group (Henricson et al., 2012; Schorr et al., 2008; Wass et al., 2008). Thus, using the CI group’s mean age might not accurately represent the age range included in all the studies. Further, norm-referenced scores can reduce the effect size (Lund, 2016), leading to low sensitivity for detecting the age effect. Lund et al. (2016) also reported that the age at testing was not significantly related to the magnitude of group differences for vocabulary knowledge. Nittrouer et al. (2018) reported no significant increases in group differences between children with CIs and their hearing peers in PA, decoding, and expressive vocabulary from second grade to sixth grade. Another study, however, reported that children with CIs started with lower scores in receptive vocabulary compared to their hearing peers, and they made enough progress yearly to reduce the gap over three years (Hayes et al., 2009). The heterogeneity between participants makes it harder to detect longitudinal changes using a cross-sectional approach. Thus, longitudinal studies are better suited to address developmental aspects of group differences between children with CIs with their hearing peers, and between children with CIs with children with HAs.

Geers (2003) found the later onset of deafness (between birth and 36 months) was associated with better reading competence. Dillon et al. (2006) also suggested that children who were not congenitally deaf performed better than children who were congenitally deaf in PA tasks. They included a wide range of ages at onset of deafness (mean ± SD: 2.3 ± 6.4, range from 0 – 36 months). However, the current meta-analysis evaluated 27 independent samples (71 effect sizes) that included a narrow range of age at onset of deafness (mean ± SD: 9.8 ± 2.7, range from 4 – 17 months). Our findings aligned with Lund et al. (2016) and Vermeulen et al. (2007), reporting that the age at onset of deafness had no significant effect on reading skills.

Several studies have reported a positive impact of age at implantation on reading skills (Archbold et al., 2008; Gallego et al., 2016; Johnson & Goswami, 2010; Mayer et al., 2016; Nittrouer et al., 2012b). Being implanted earlier in life was associated with better PA and linguistic comprehension (Nittrouer et al., 2012b). Despite many benefits of early implantation, children with CIs, regardless of age at implantation, continue to have difficulties with grammatical comprehension than their hearing peers (Gallego et al., 2016). Especially when cognitive processing demands increased, children with CIs performed more poorly than their hearing peers, which kept them from reaching a normalized reading comprehension level in most cases (Gallego et al., 2016). All those studies divided children with CIs into early-implantation and late-implantation groups. However, the current meta-analysis used age at implantation as a continuous variable (mean ± SD: 31.8 ± 18.1, range: 14.7 – 99.5 months) and might not be sensitive to capturing the effect of age at implantation on group differences in reading skills between children with CIs and their hearing peers. Our findings are aligned with some studies reporting that the age at implantation did not significantly affect PA, vocabulary, and decoding (Ambrose, 2009; Lund, 2016; Wass et al., 2008) and a recent review article (Mayer & Trezek, 2018).

The percent of children who used a unilateral CI in the study cohort was the only significant moderator. Findings from the current meta-analysis demonstrated that a higher percentage of children with a unilateral CI in the study cohort was associated with more substantial group differences in reading skills between the CI and TH groups, suggesting bilateral implantation has more benefits for reading skills than unilateral implantation. Bilateral implantation has been reported to have a positive effect on PA, linguistic comprehension, receptive vocabulary, and rate of vocabulary development, where children with bilateral CIs outperformed children with a unilateral CI (Boons et al., 2012; Caselli et al., 2012; Guerzoni et al., 2020; Sarant et al., 2014; Sparreboom et al., 2015). Additionally, children with a unilateral CI and a contralateral HA performed more poorly in reading comprehension tasks than children with bilateral CIs (Guerzoni et al., 2020). The benefits of bilateral implantation can be affected by several factors, such as whether the implants were received simultaneously or sequentially, the inter-implant interval length if they were sequential, and the age when the first and second implants were received. Simultaneous bilateral implantation and short inter-implant intervals were significantly associated with better speech perception outcomes (Gordon & Papsin, 2009). Longer inter-implant intervals increased the chance of not using the second implant regularly to maximize the benefits of bilateral implantation (Low et al., 2020). In addition, children with unilateral CI and HA in the other better ear who experienced early bimodal stimulation had better PA skills than children with bilateral Cis (Nittrouer et al., 2018). Children with bilateral CIs or children who experience bimodal stimulation had a better chance of developing good speech perception skills (de Raeve et al., 2015).

### Limitations and Future Directions

There are several limitations to the current meta-analysis. First, the limited amount of information available about socioeconomic status, home literacy environment, and nonverbal cognition constrain the moderator analysis in the present study. These factors, including socioeconomic status, home literacy environment, and nonverbal cognition, have been related to reading development for both CI and TH groups (Traxler, 2017). Group differences in reading skills may result, wholly or partly, from different early language and reading experiences at home, and from how children were taught to read in school. Future work may consider these factors in a correlational meta-analysis. In addition, future studies need to include more detailed information about the characteristics of hearing loss, environmental factors, and early interventions received (e.g., whether sign language was used before cochlear implantation). These factors are essential to resolve the discrepancy among studies in the current literature. Second, both published and unpublished studies were included in the present study to minimize publication bias. The asymmetric funnel plot for comparison between CI and TH groups indicated the presence of small-study effects. Although Rosenthal’s Fail-safe N suggested that it is unlikely that the actual group difference is zero, the magnitude of standardized mean difference estimates might be overstated for comparison between CI and TH groups. Third, the number of studies included in the meta-analysis of group differences between children with CIs and children with HAs was relatively small. Very few studies compared reading skills between the CI and HA groups. Moreover, not all studies reported information regarding the characteristics of hearing loss for each group. Thus, there was not enough data to conduct the same moderator analysis to compare the CI and HA groups. Finally, given the limited research available, 21 articles comparing children with CIs to their hearing peers and six articles comparing children with CIs to children with HAs were carried out in language (i.e., Swedish, French, German, Dutch, Spanish, Farsi, Turkish, Persian, Hindi, Korean, Chinese) other than English were not excluded in the current meta-analysis (see Supplementary Table A.2), which might introduce more variance in effect sizes. Chinese is a logographic writing system that is different from English using an alphabetical system. Better PA in Chinese readers has been significantly correlated to better Chinese word recognition (Huang & Hanley, 1995). Within 41 articles comparing children with CIs to their hearing peers, one study (Tse & So, 2012) conducted PA tasks in Cantonese (effect size range from .008 — -.538, see Supplementary Figure A.1) and is unlikely to affect the overall effect size for PA (g = -1.62).

Despite these limitations, the current meta-analysis highlights the positive shift in reading skills for the newer generation of children with CIs, even though the use of CIs did not normalize PA, vocabulary, decoding, fluency, and reading comprehension skills for children with CIs. Since most children with CIs in the U.S. are mainstreamed (Vatalaro et al., 2018), reading challenges in children with CIs might be overlooked. Educators need to be mindful that some children with CIs may need additional support in the early intervention and educational settings. Future studies can focus on designing appropriate reading assessments and evaluating the effects of code-based or meaning-based intervention for children who are DHH.

## Data Availability

Data and codes are available by request.

## Acknowledgments

This work was supported by the National Institute on Deafness and Other Communication Disorders [5R21DC018110, 2019]; the Barkley Trust, Nebraska Tobacco Settlement Biomedical Research Development, College of Education and Human Sciences, and the Office of Research and Economic Development at University of Nebraska-Lincoln (UNL). The content of this paper is solely the responsibility of the authors and does not necessarily represent the official views of the NIH. The undergraduate research assistant (Makayla J. Gill) was supported by the UNL Undergraduate Creative Activities and Research Experience (UCARE) program funded in part by gifts from the Pepsi Quasi Endowment and Union Bank & Trust. Many thanks to Dr. Michelle L. Hughes for providing her constructive comments. Y.W. conceptualized research ideas, performed meta-analyses, and wrote the main paper. F.S. coded all the articles, performed initial analysis, and drafted part of the paper. K.L. provided statistical analysis, wrote R codes to analyze and valid results, and edited the statistical methodology part in the paper. M.J.G coded 80% of the studies and edited the paper. J.H. reviewed and edited the paper. All authors commented on the manuscripts before the submission. All authors declare that there is no conflict of interest. The data that support the findings of this study are available by request to the corresponding author.

*\* marked articles that are included in the quantitative meta-analysis and a full list of all articles that included in the meta-analysis can be found in the Supplementary Document.*

## Appendix A. Supplementary Tables

**Supplementary Table A.1.** Study Characteristics

| Group Comparison |  | CI vs. TH |  | CI vs. HA |
| --- | --- | --- | --- | --- |
| Study Characteristic | # of articles | Percentage (%) | # of articles | Percentage (%) |
| **Publication Year** |  |  |  |  |
| 2002-2005 | 2 | 5 | 1 | 7 |
| 2006-2010 | 7 | 17 | 3 | 20 |
| 2011-2015 | 20 | 49 | 9 | 60 |
| 2016-2019 | 12 | 29 | 2 | 13 |
| **Language** |  |  |  |  |
| English | 20 | 49 | 9 | 60 |
| Swedish | 5 | 12 | 3 | 20 |
| Farsi | 4 | 11 | 1 | 7 |
| Flemish | N/A | N/A | 1 | 7 |
| Korean | 2 | 5 | N/A | N/A |
| Spanish | 2 | 5 | N/A | N/A |
| Turkish | 2 | 5 | N/A | N/A |
| Mandarin | 2 | 5 | 1 | 7 |
| Dutch | 1 | 2 | N/A | N/A |
| Hindi | 1 | 2 | N/A | N/A |
| German | 1 | 2 | N/A | N/A |
| Italian | 1 | 2 | N/A | N/A |
| **Publication** |  |  |  |  |
| Journal | 38 | 93 | 14 | 93 |
| Thesis | 3 | 7 | 1 | 7 |
|  | # of effects | Percentage (%) | # of effects | Percentage (%) |
| **Sample Size of Children with CIs** |  |  |  |  |
| <=10 | 20 | 15 | 11 | 19 |
| 11-30 | 91 | 68 | 43 | 73 |
| 31-50 | 14 | 10 | 5 | 8 |
| 51-70 | 8 | 6 |  |  |
| >=71 | 1 | 1 |  |  |

**Supplementary Table A.2.**
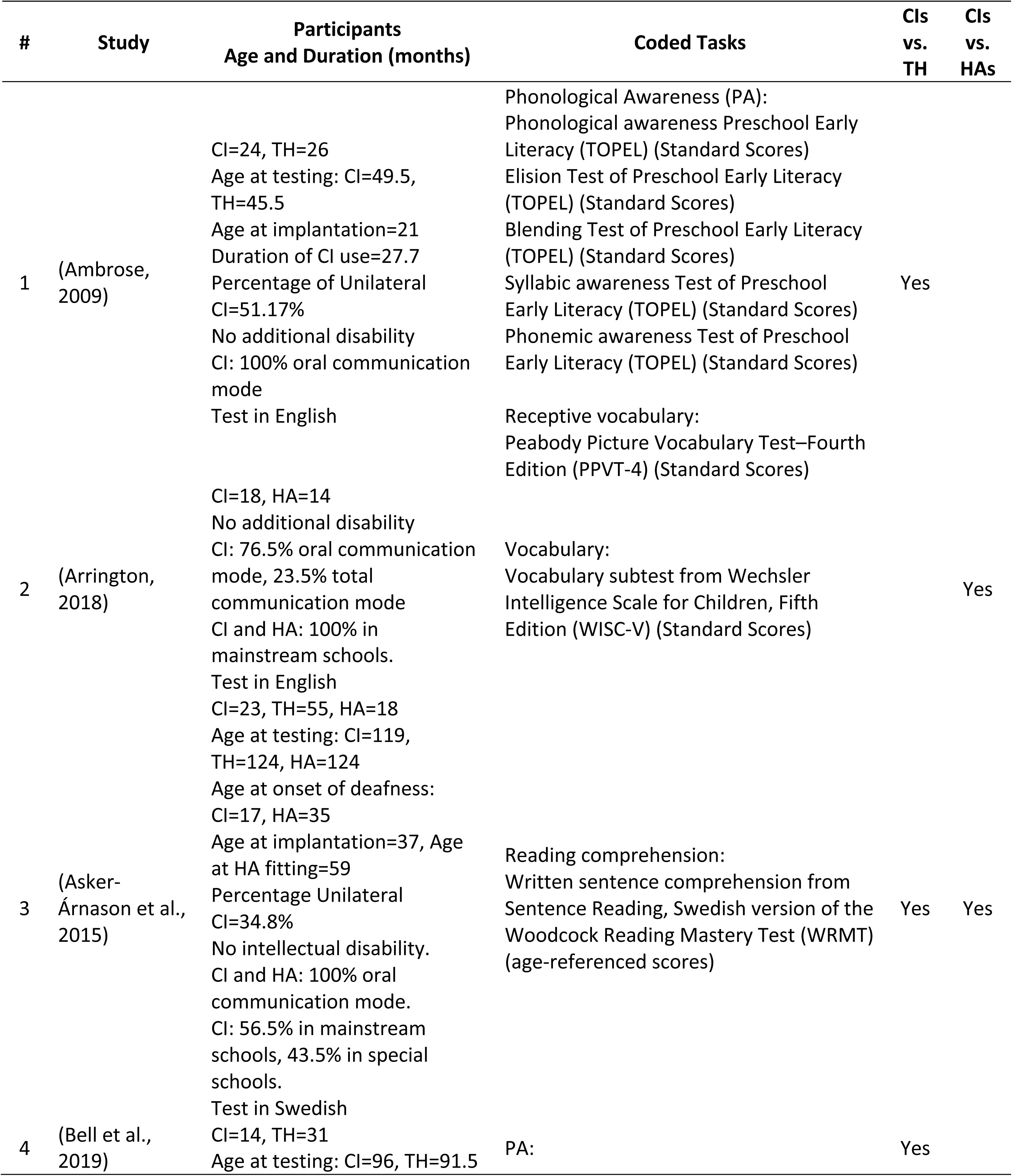
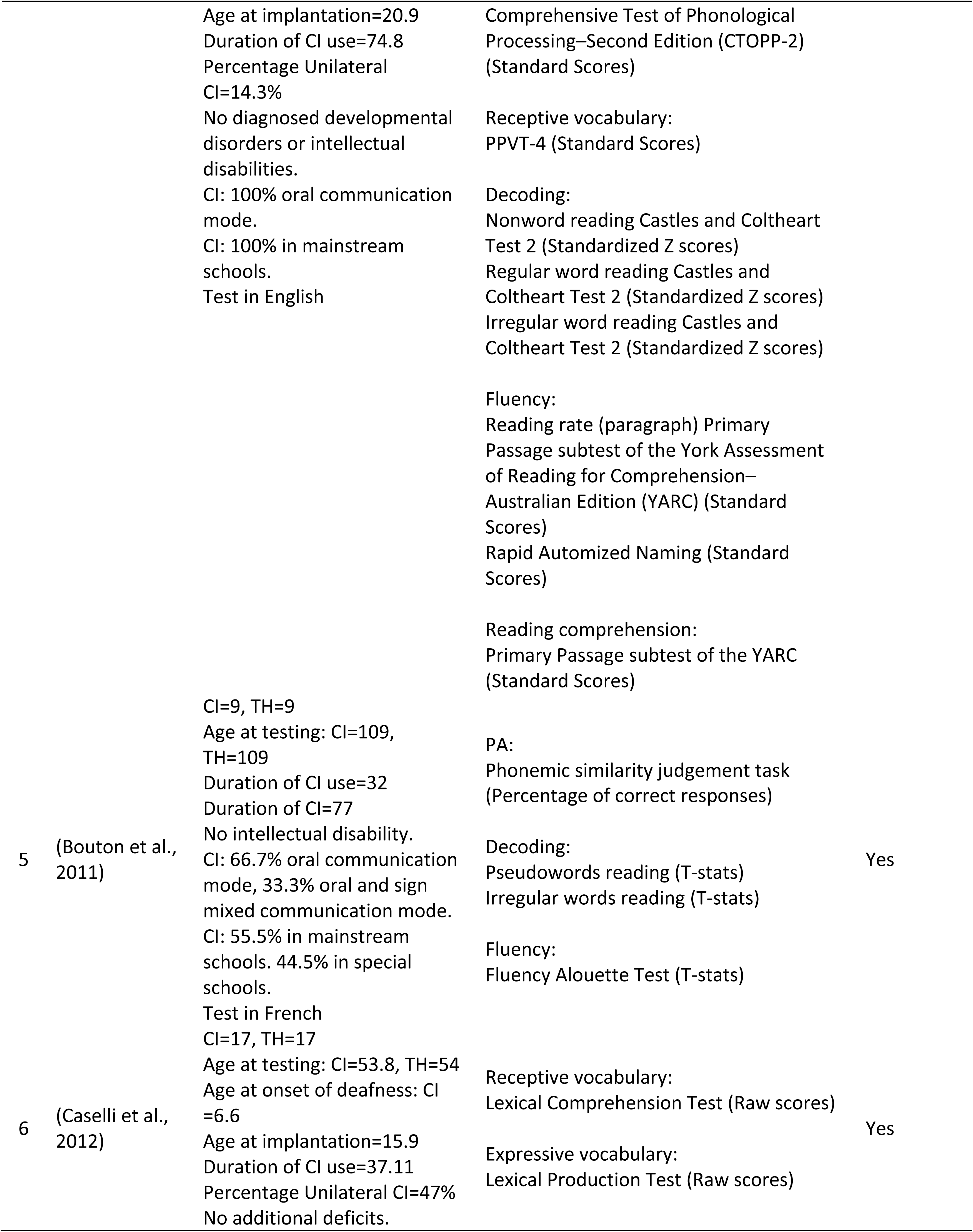
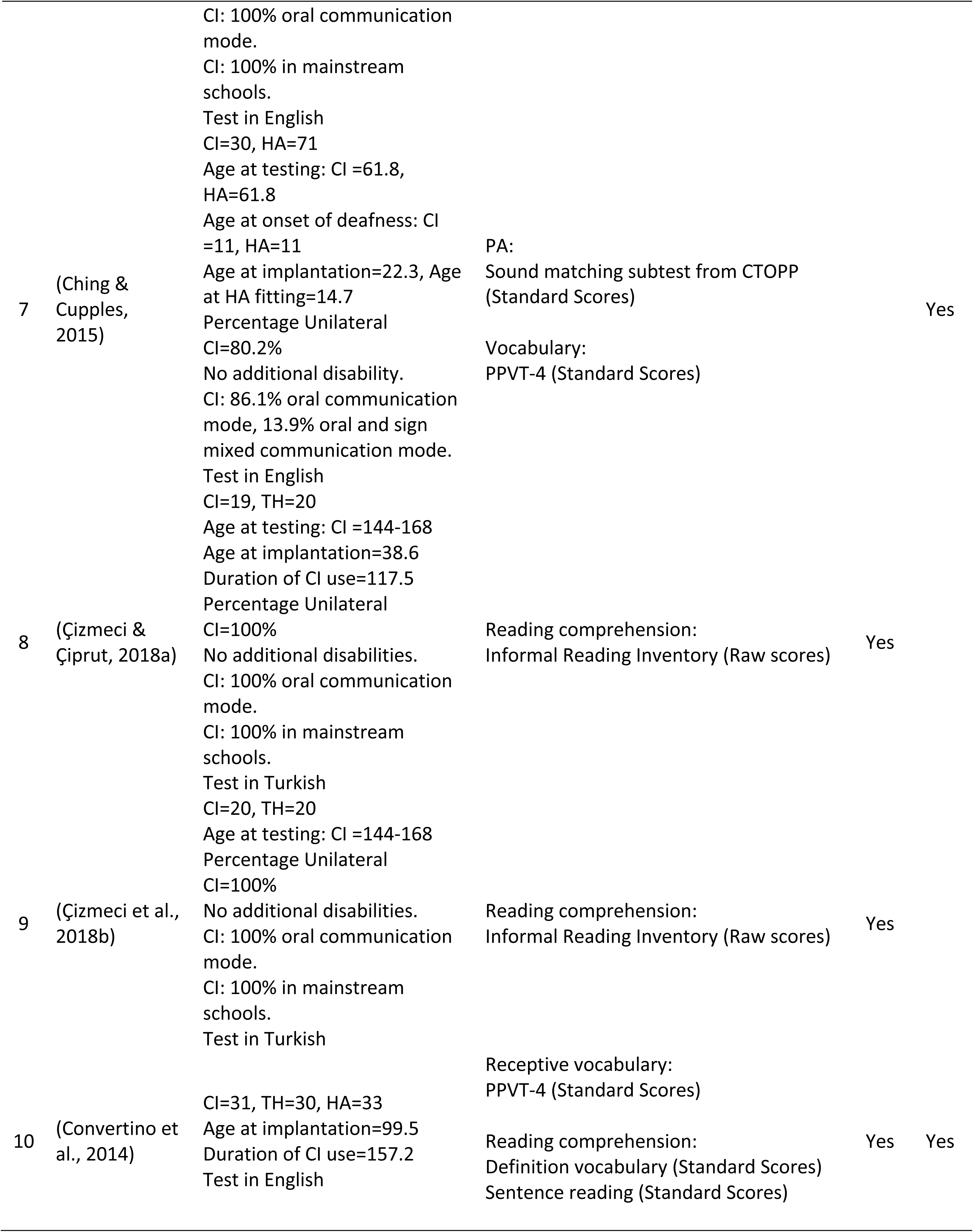
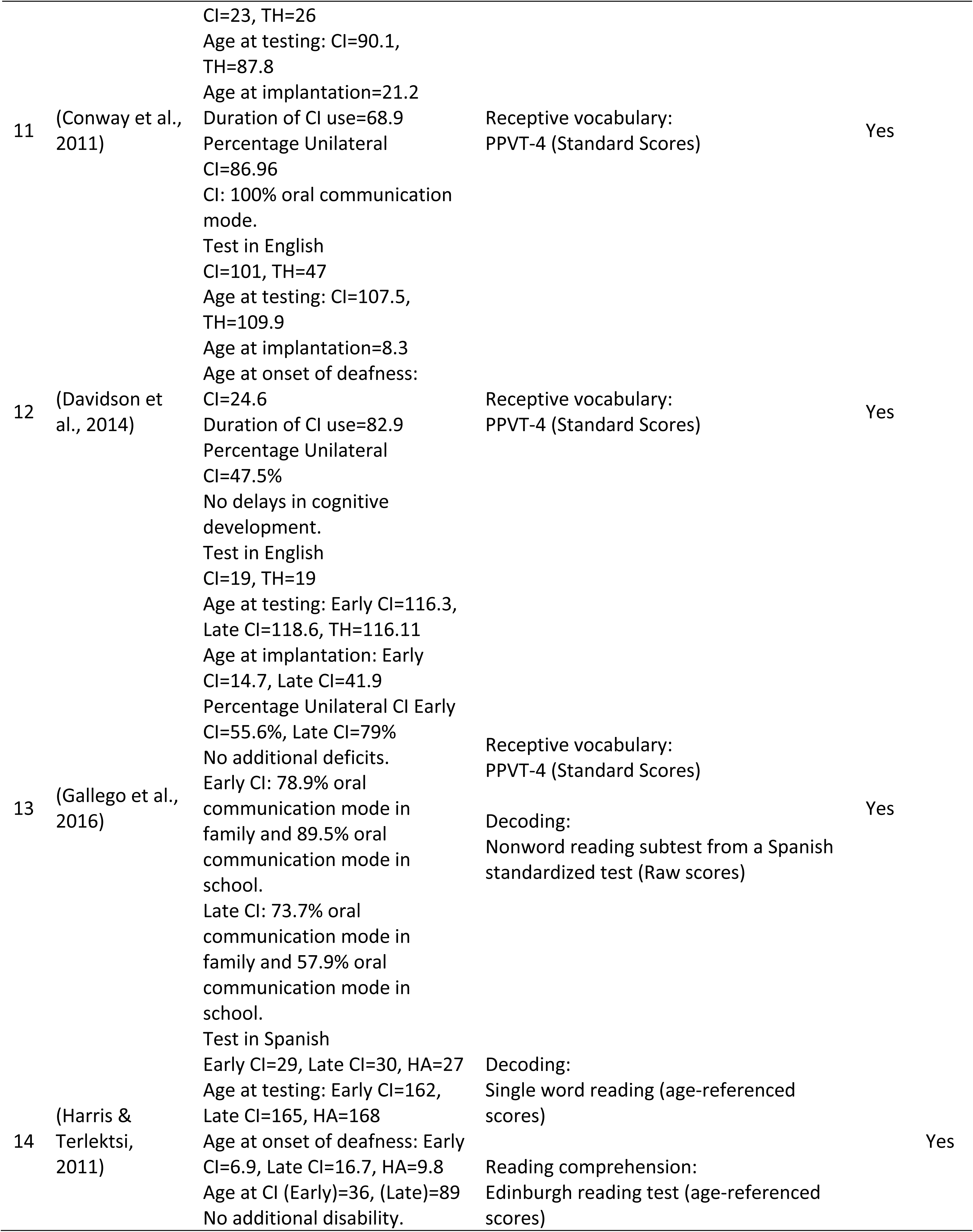
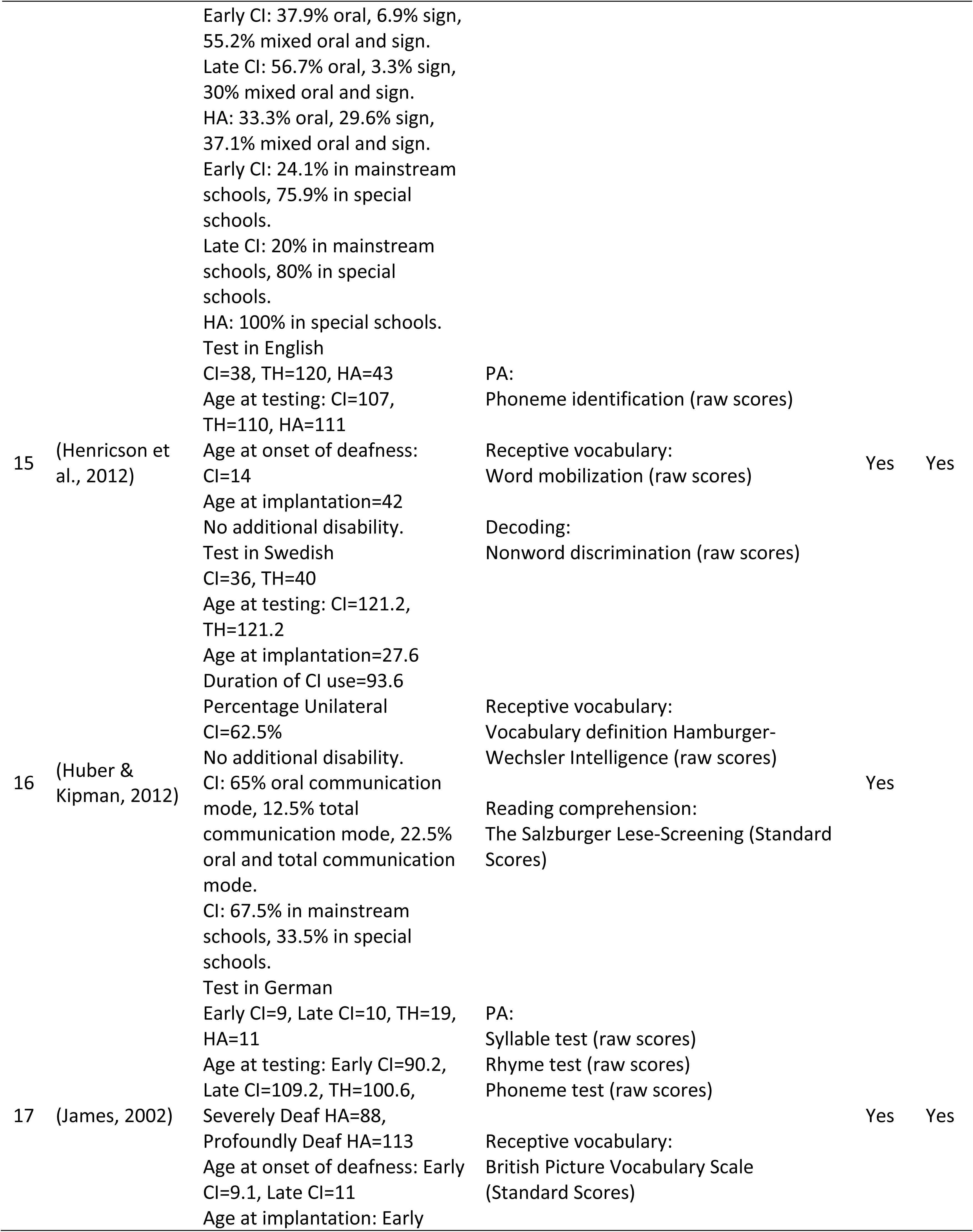
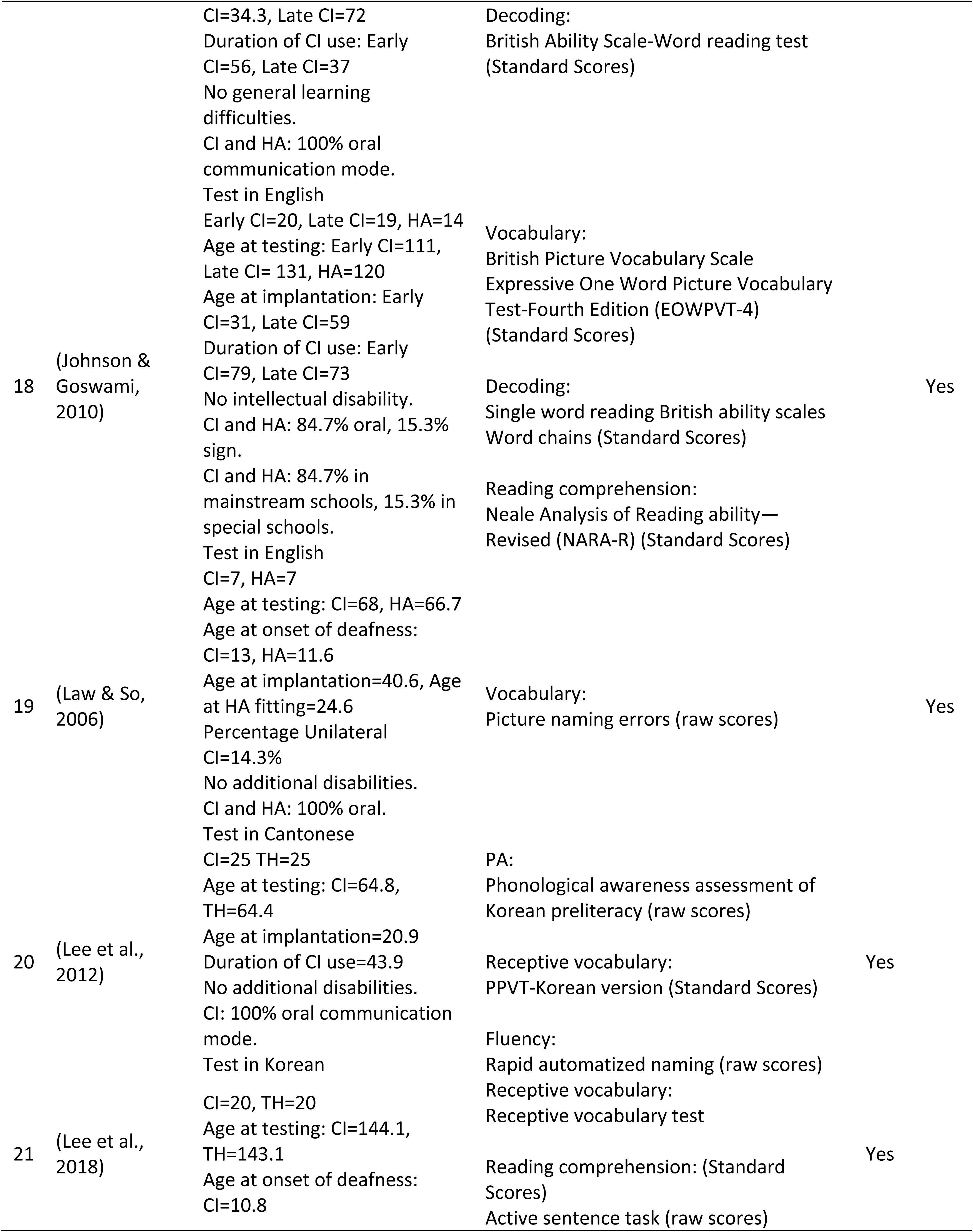
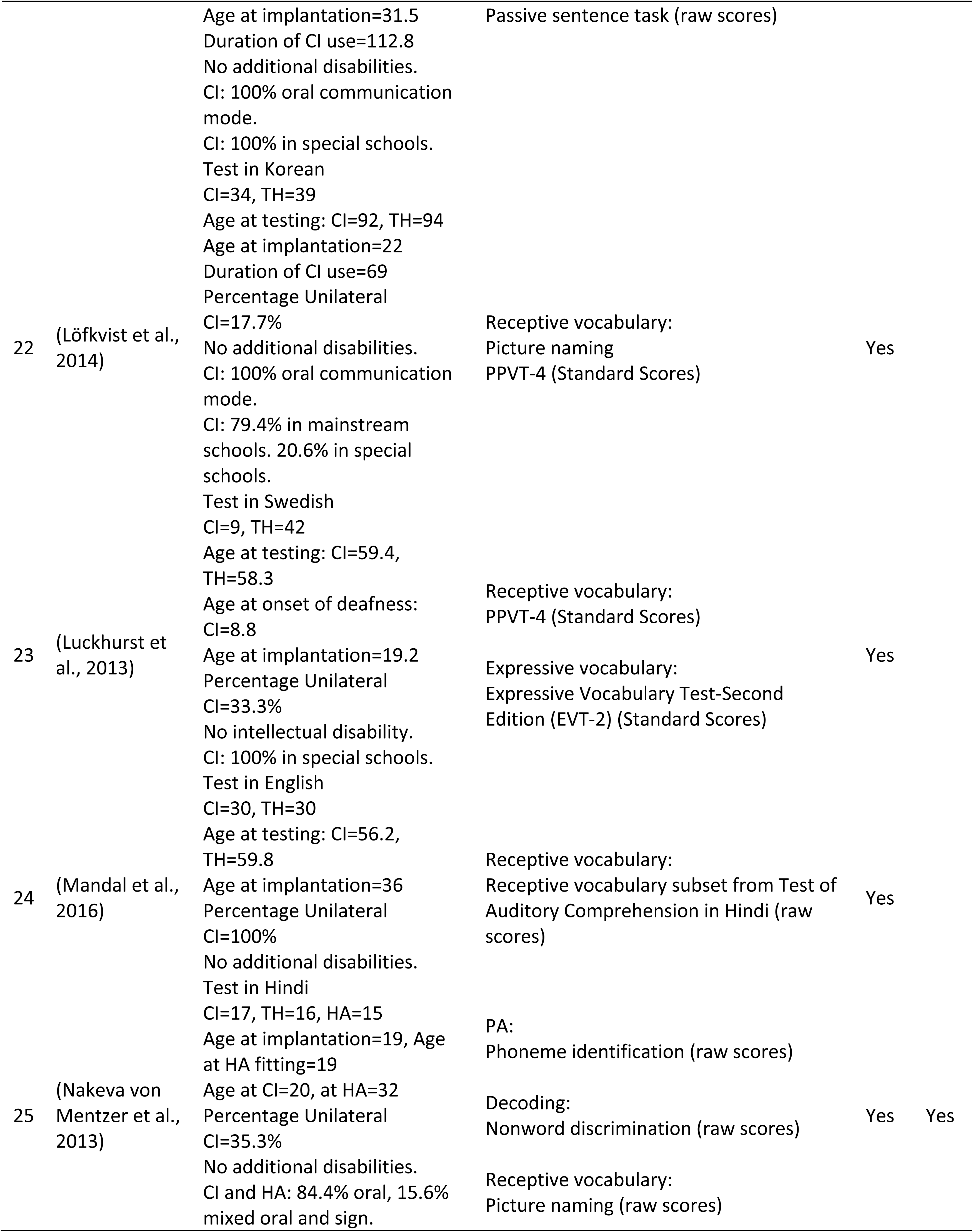
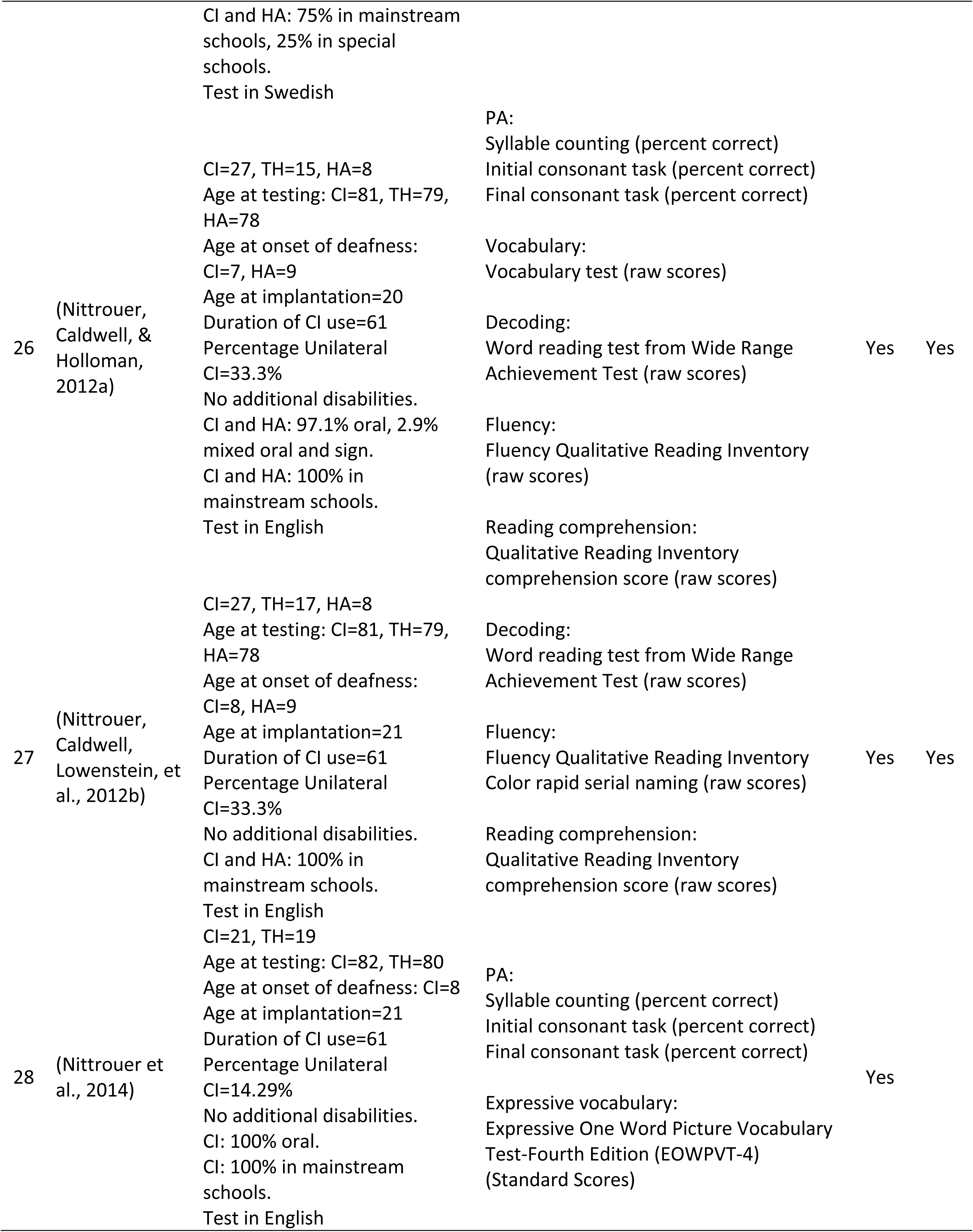
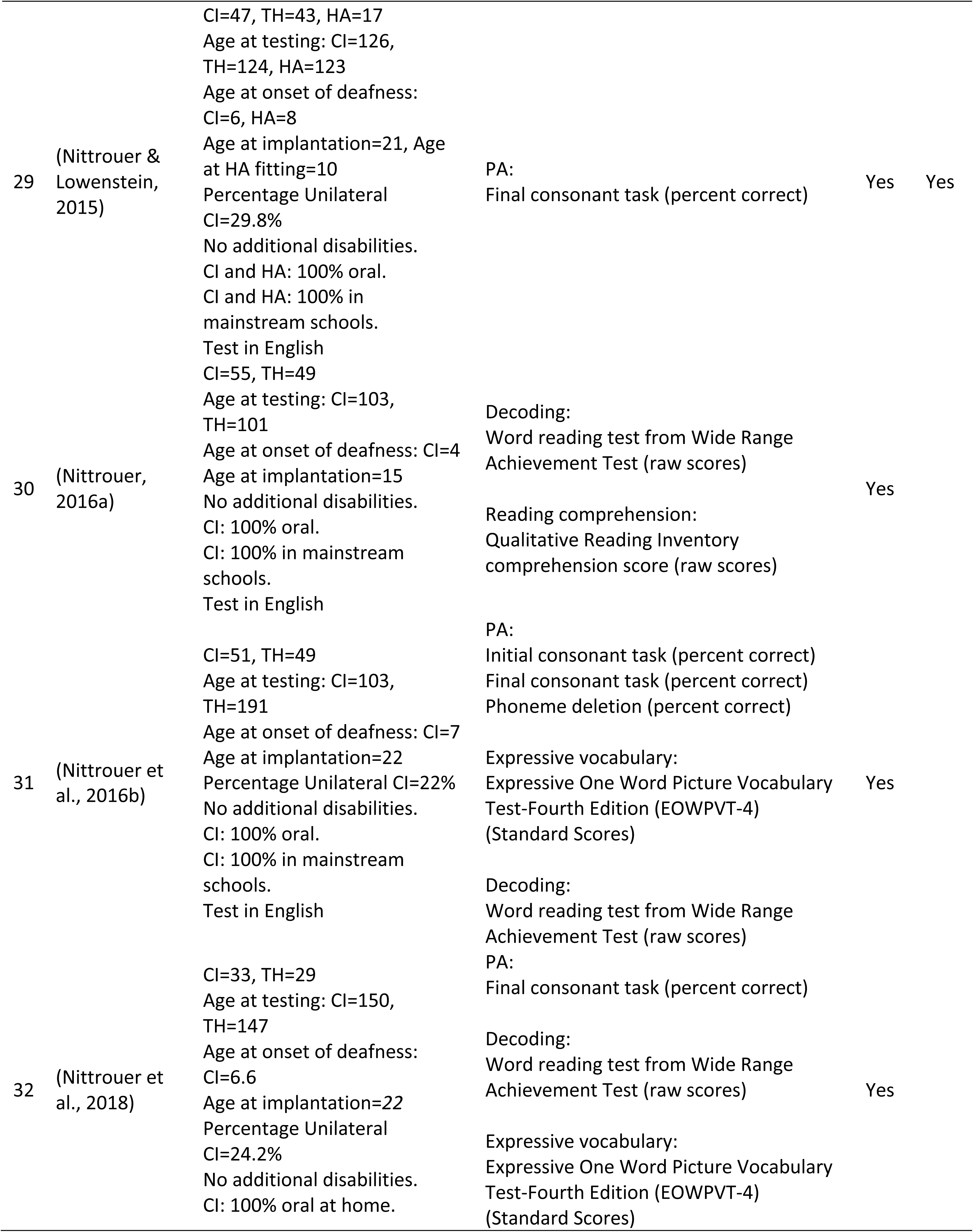
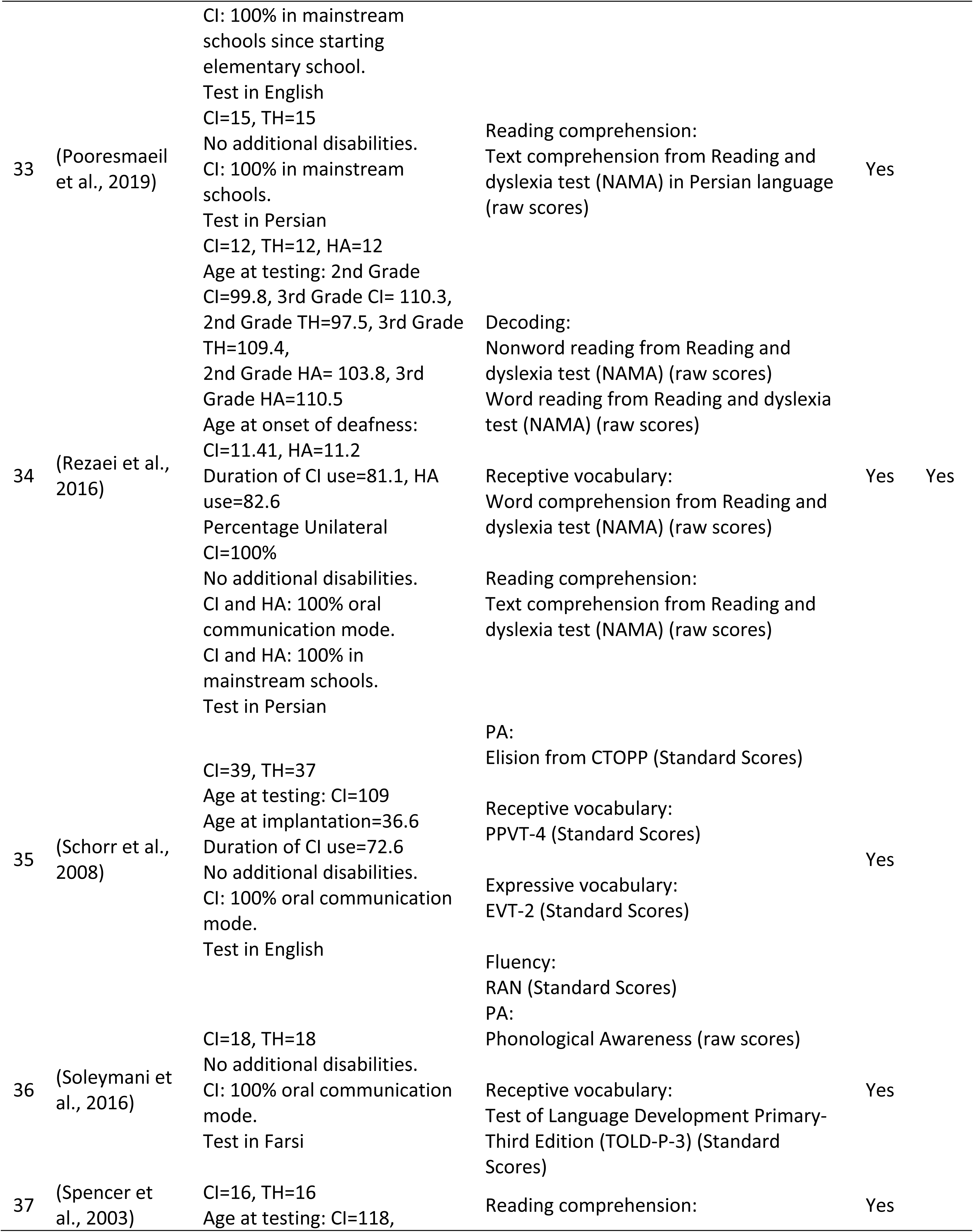
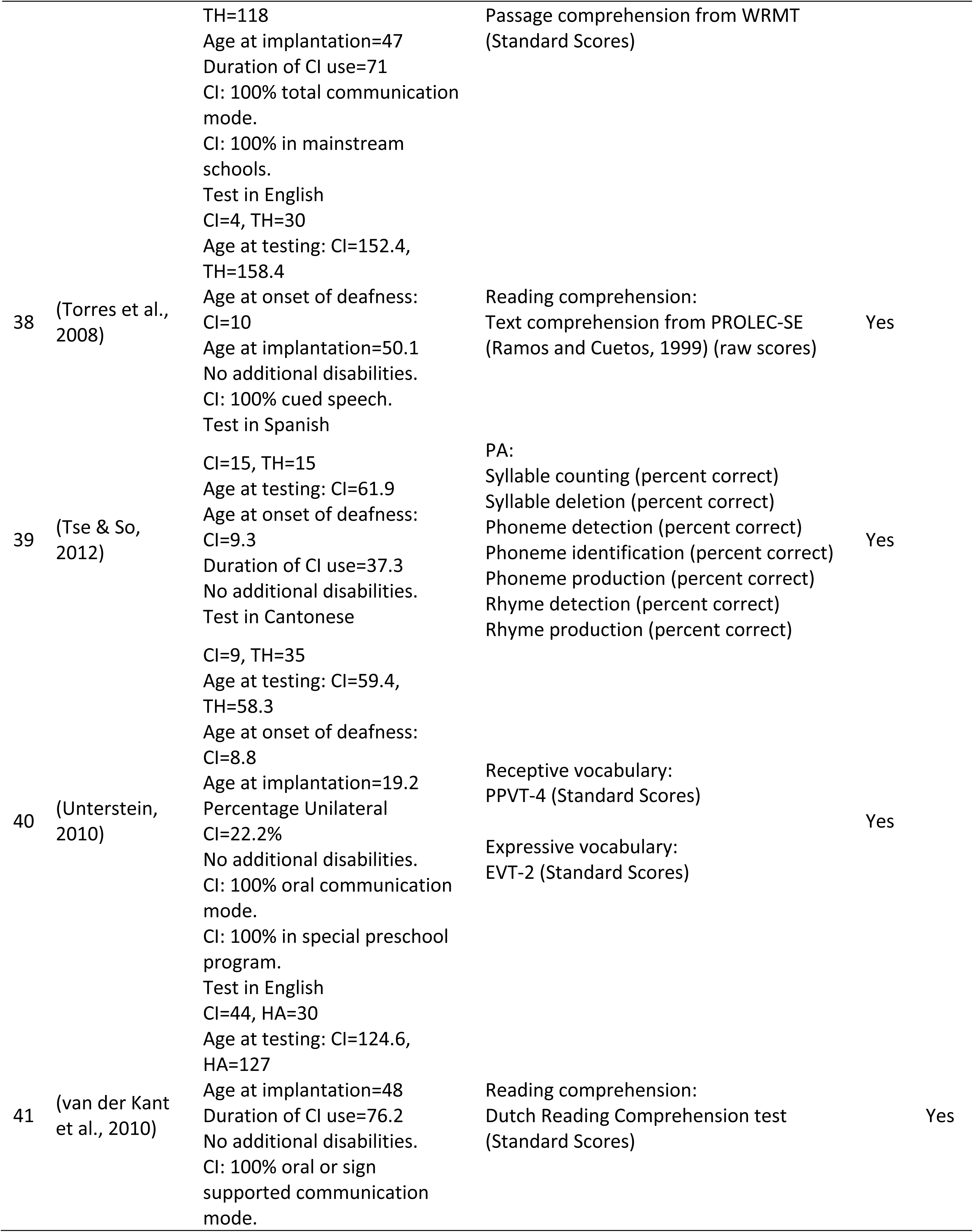
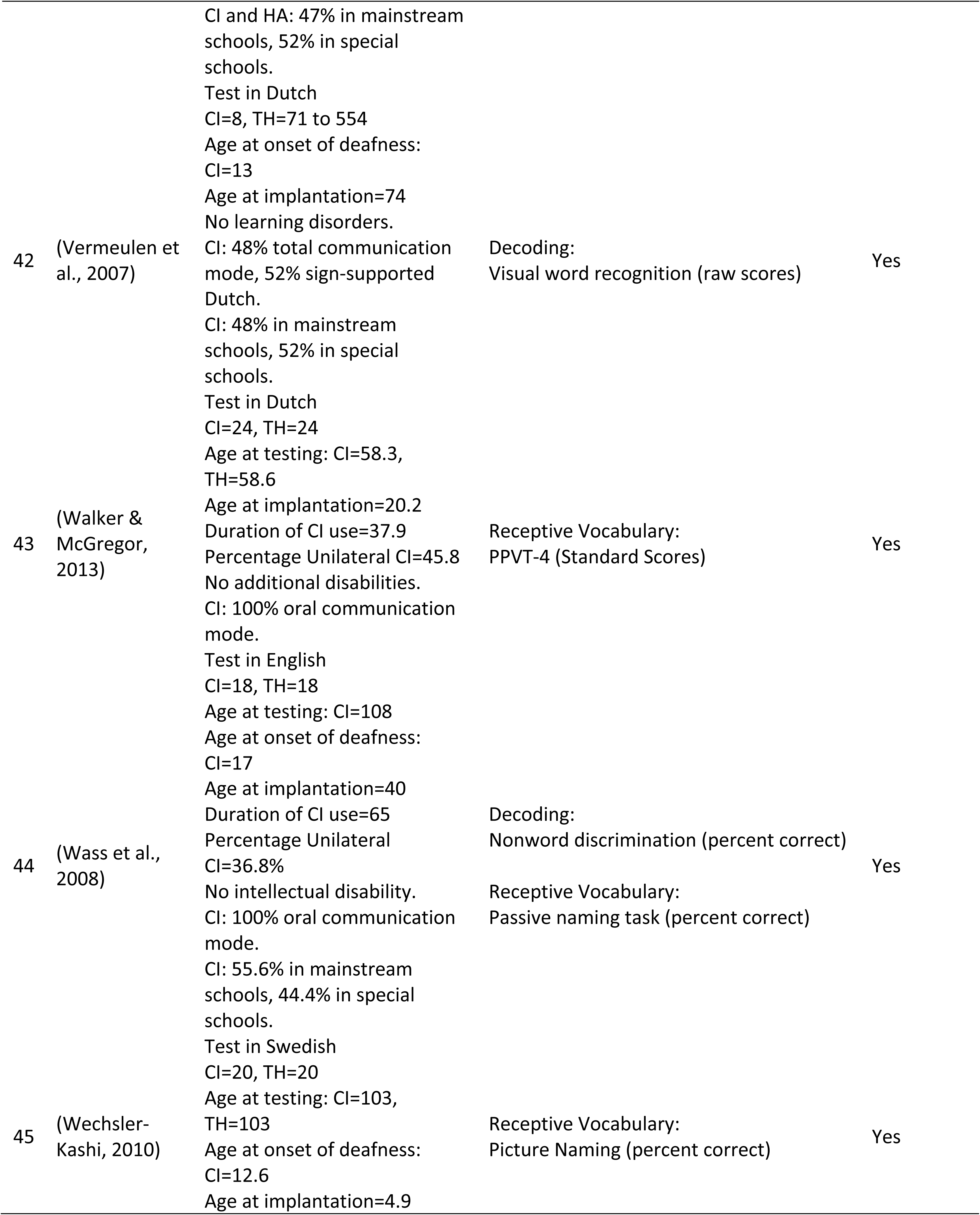
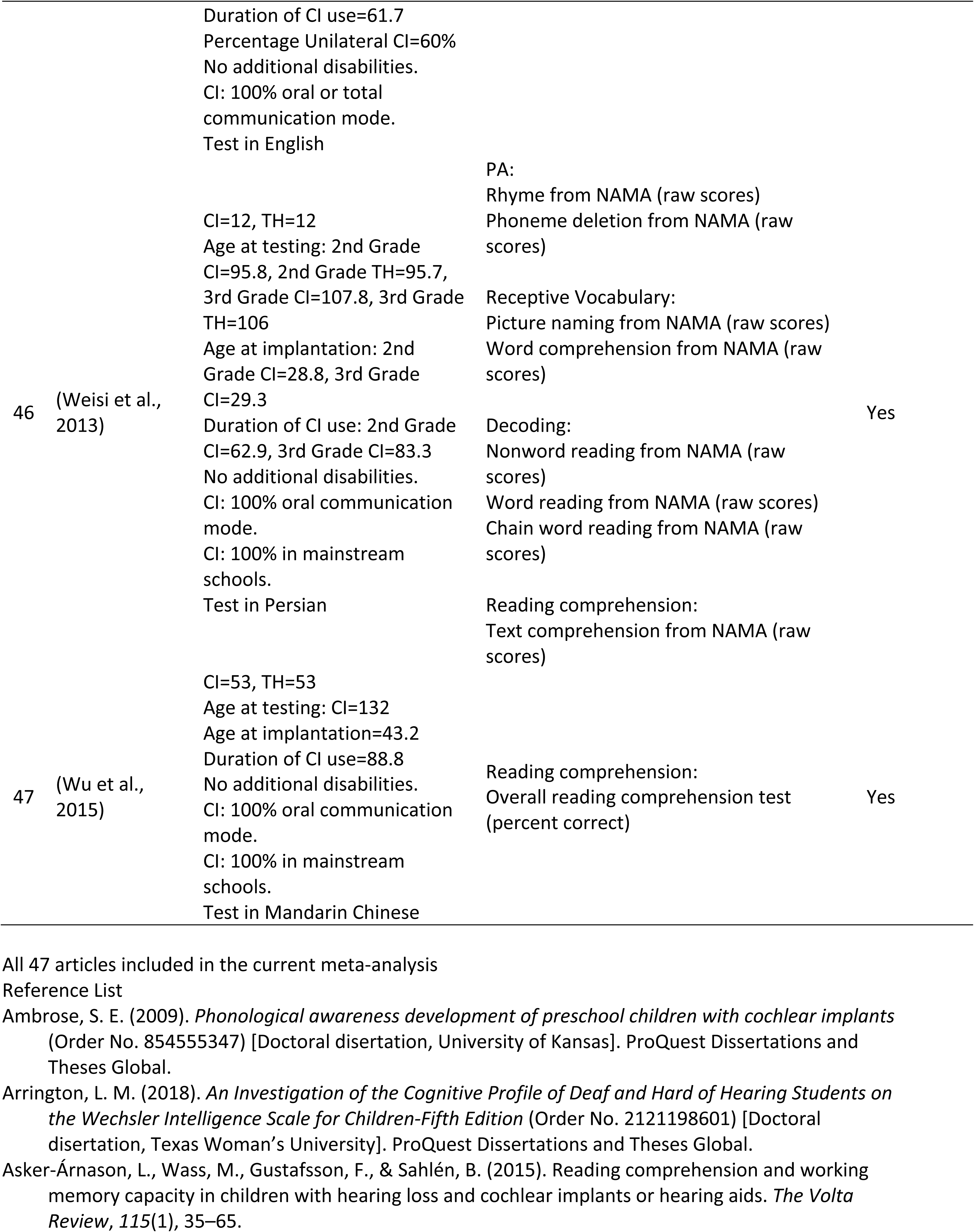
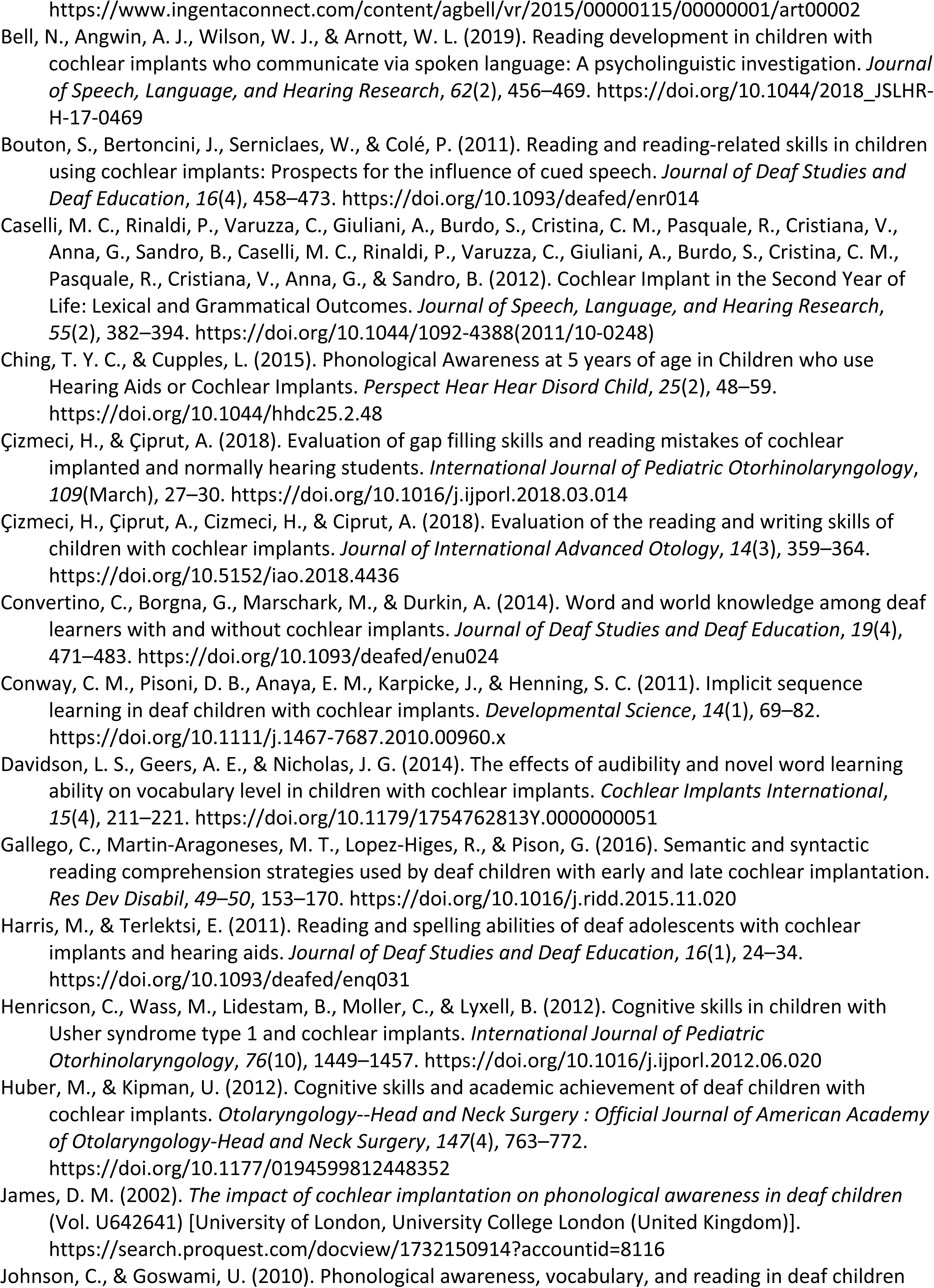
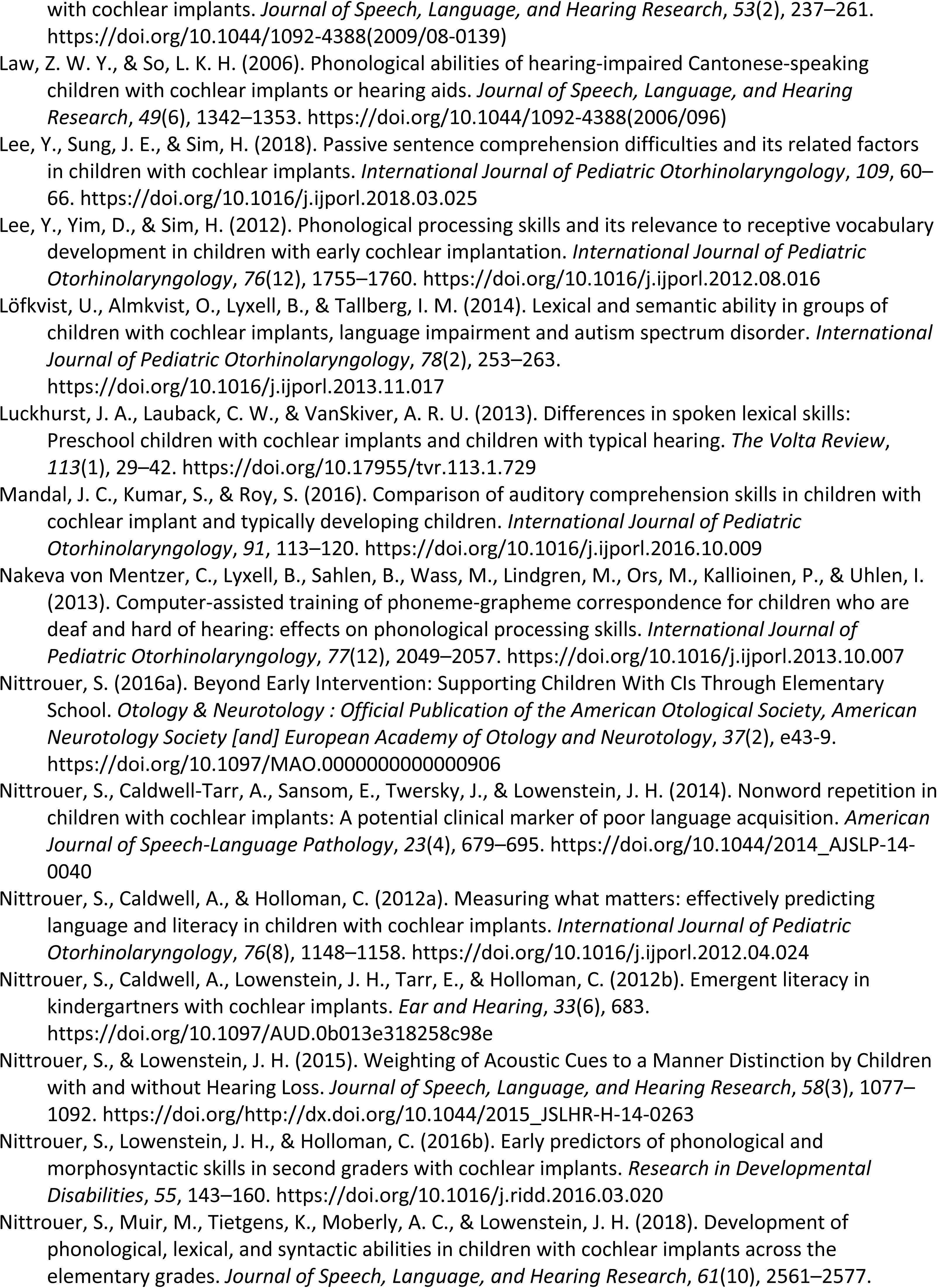
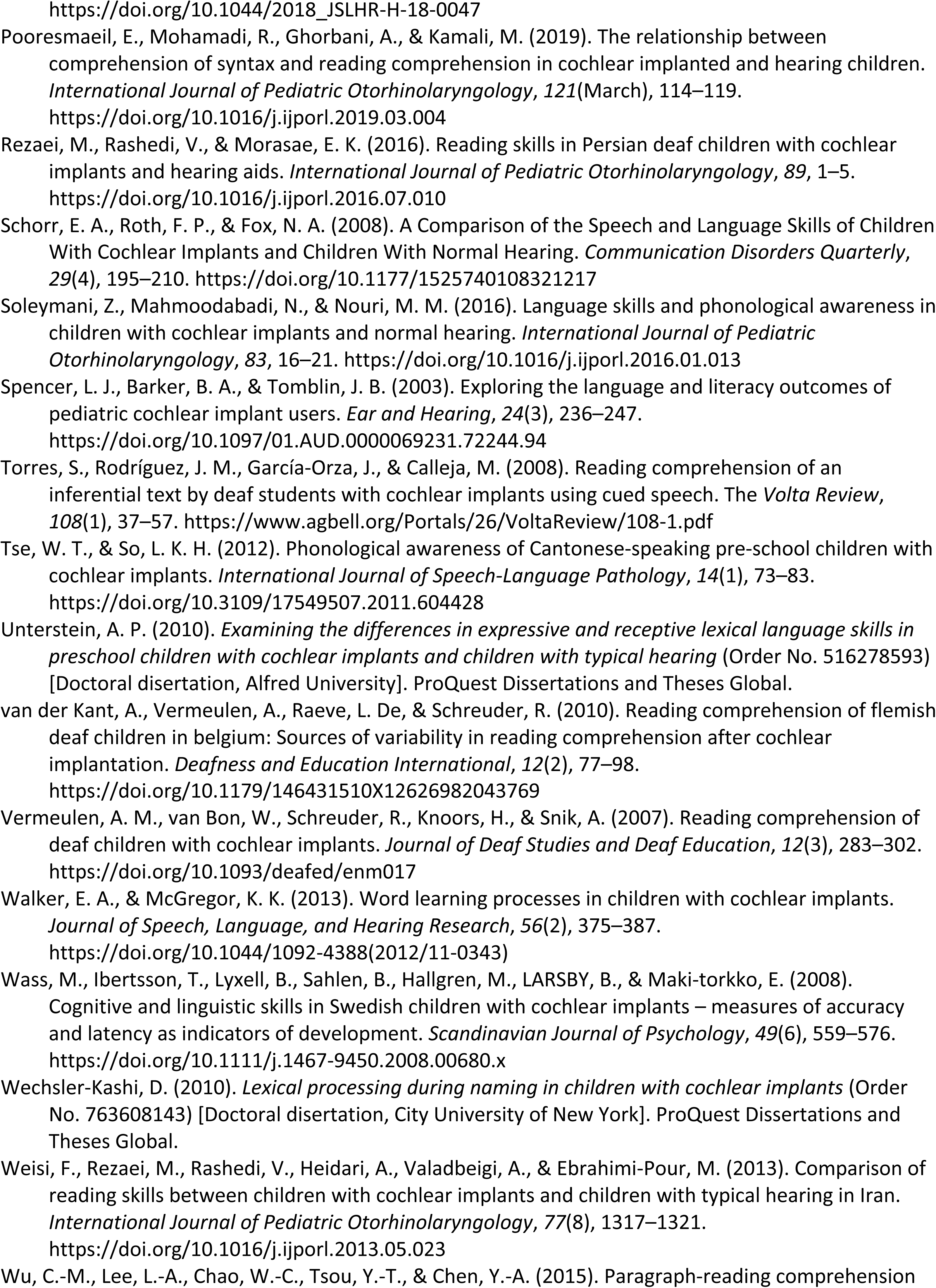
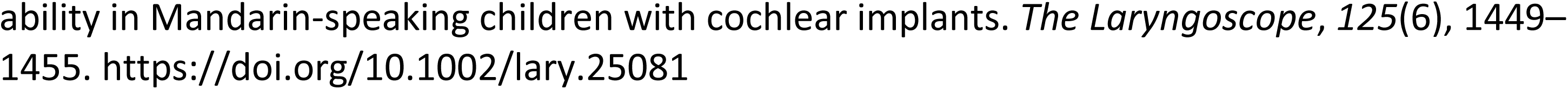
All articles included in the current study

**Supplementary Table A.3.**
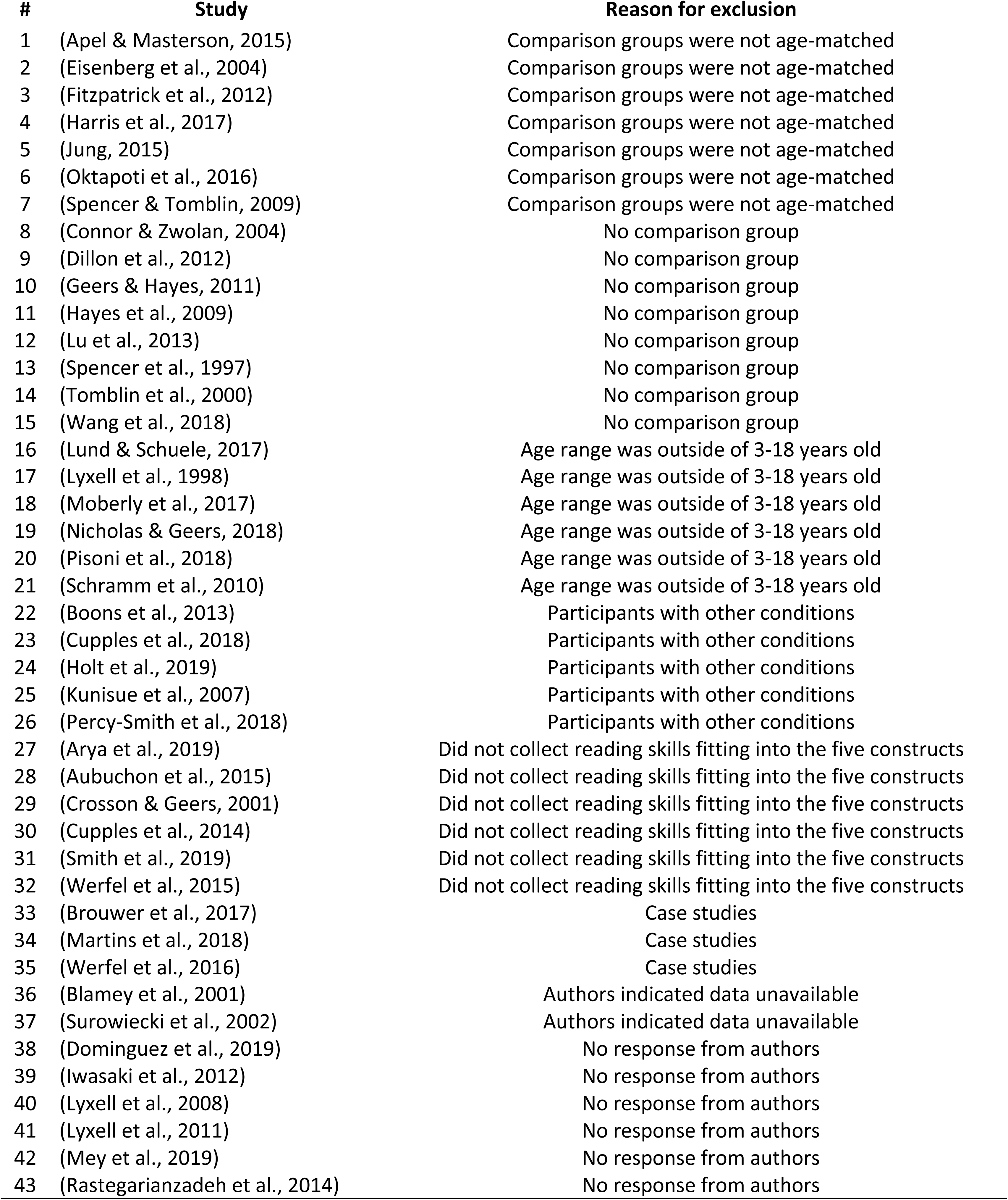
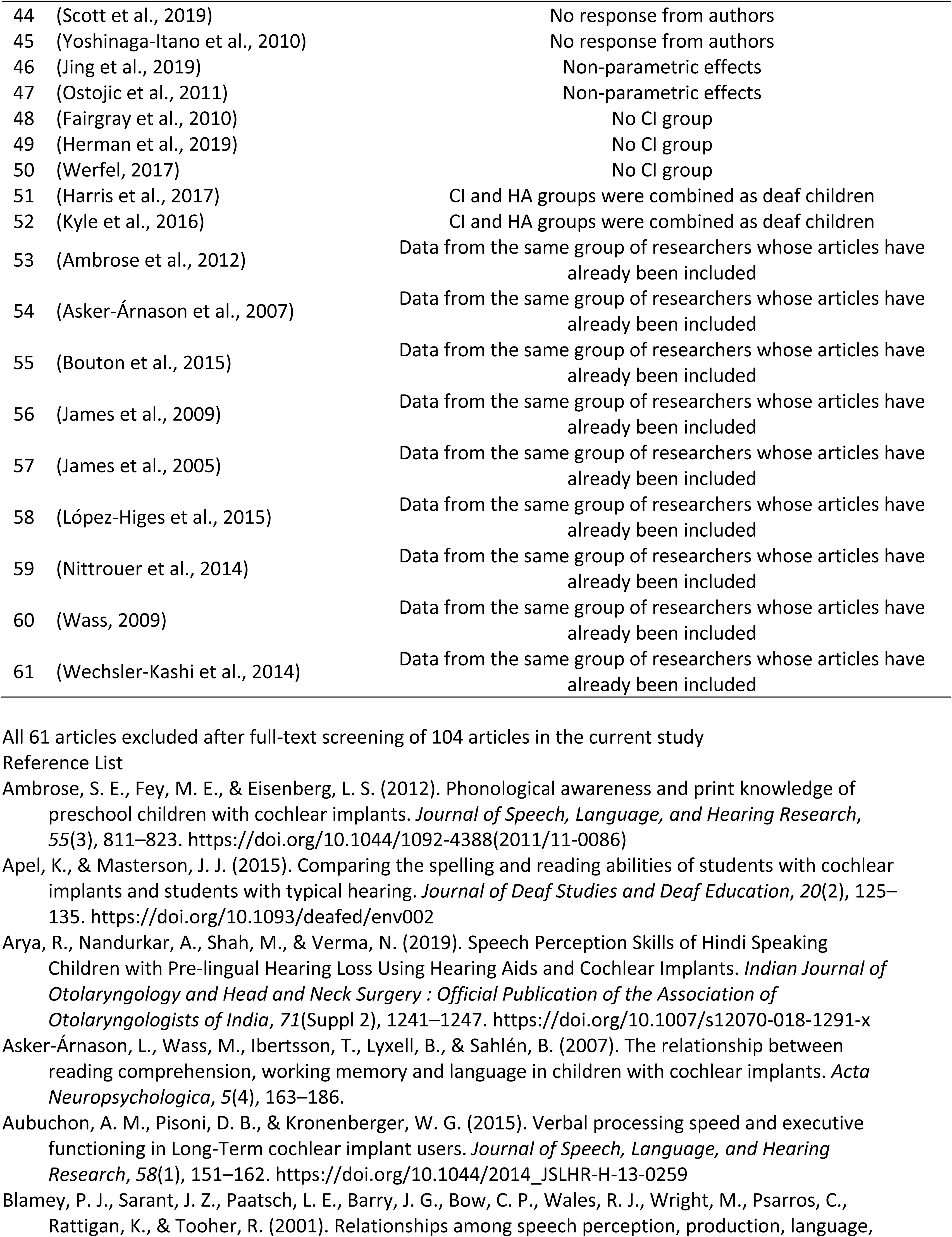
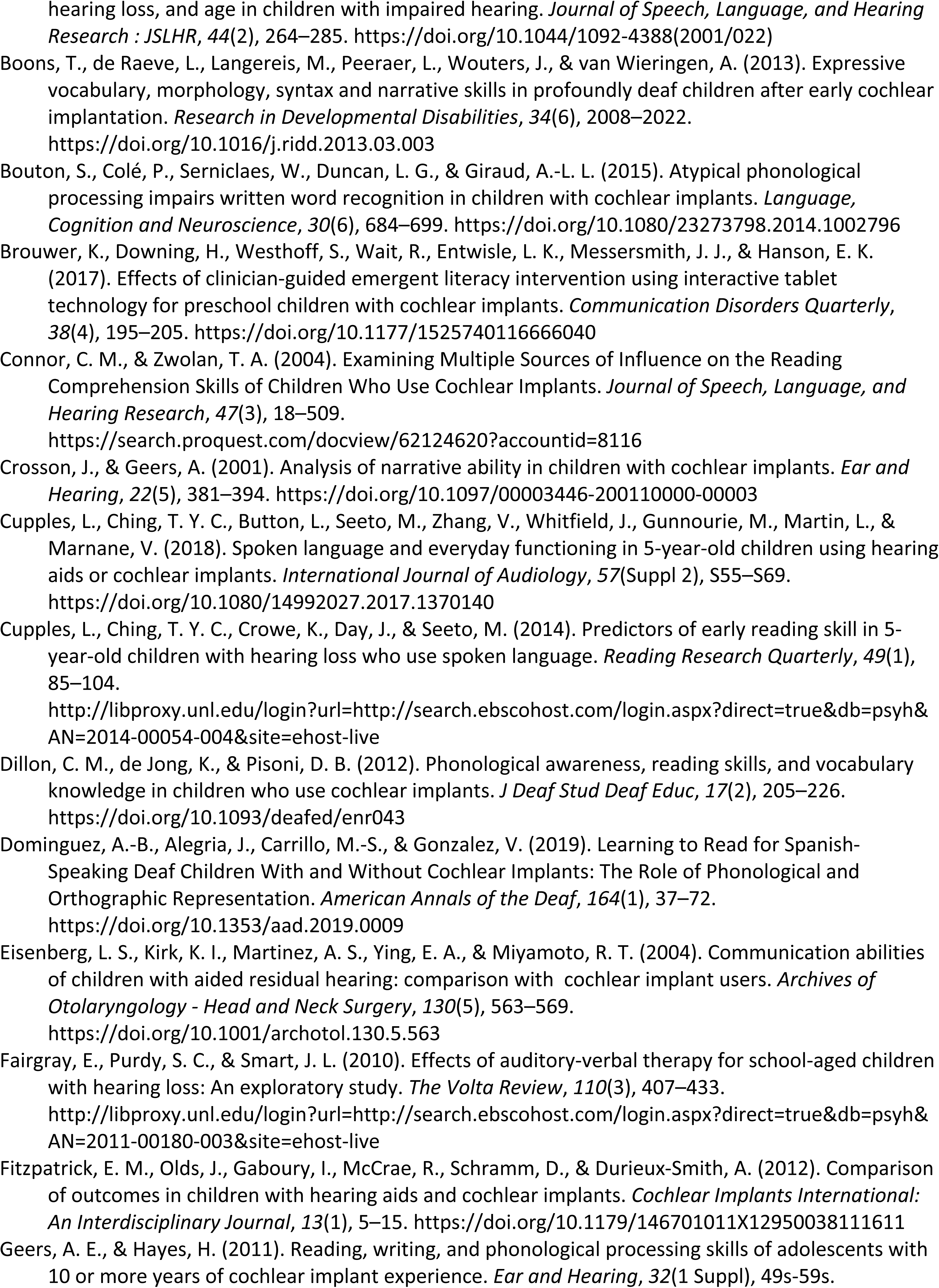
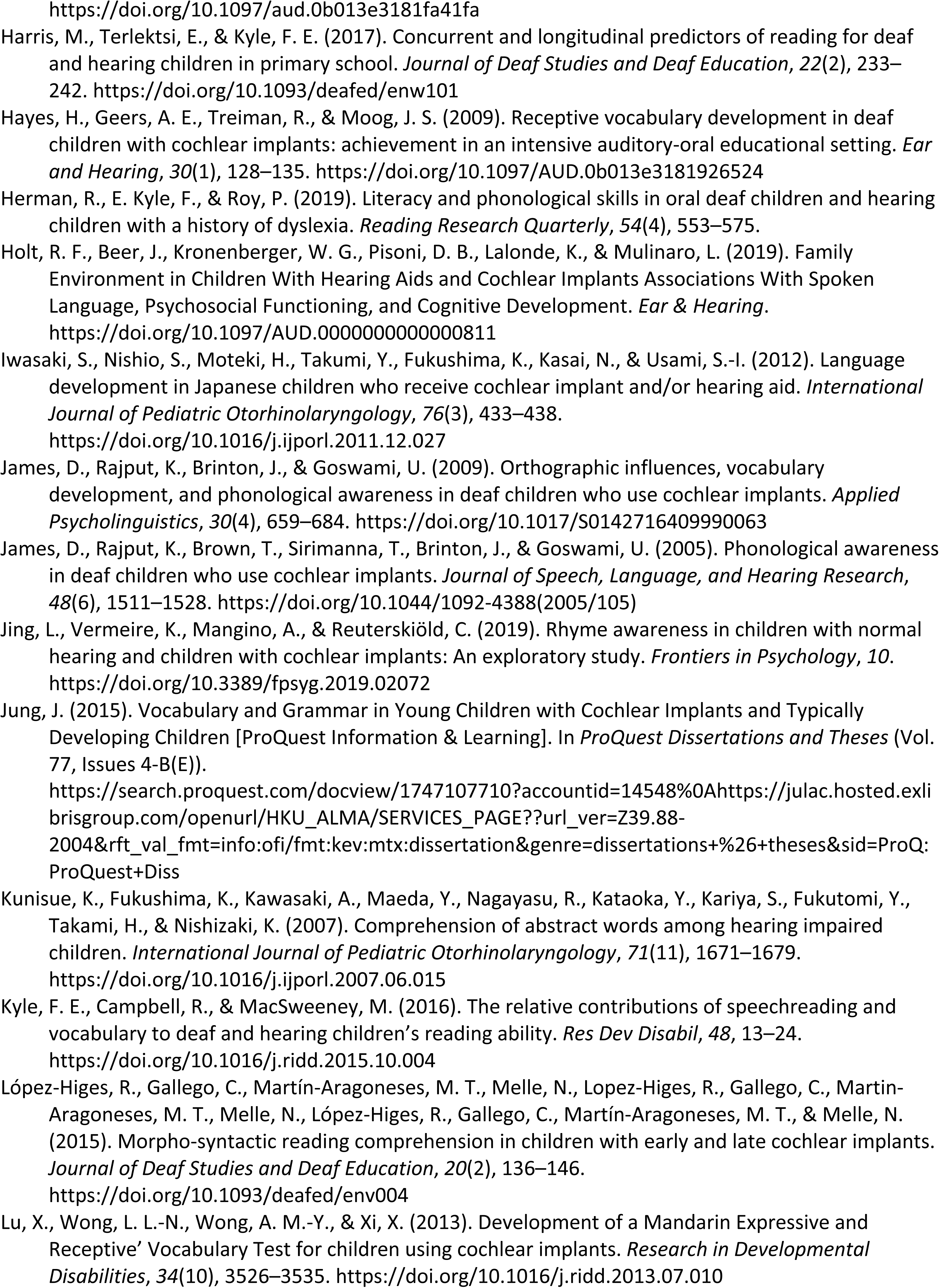
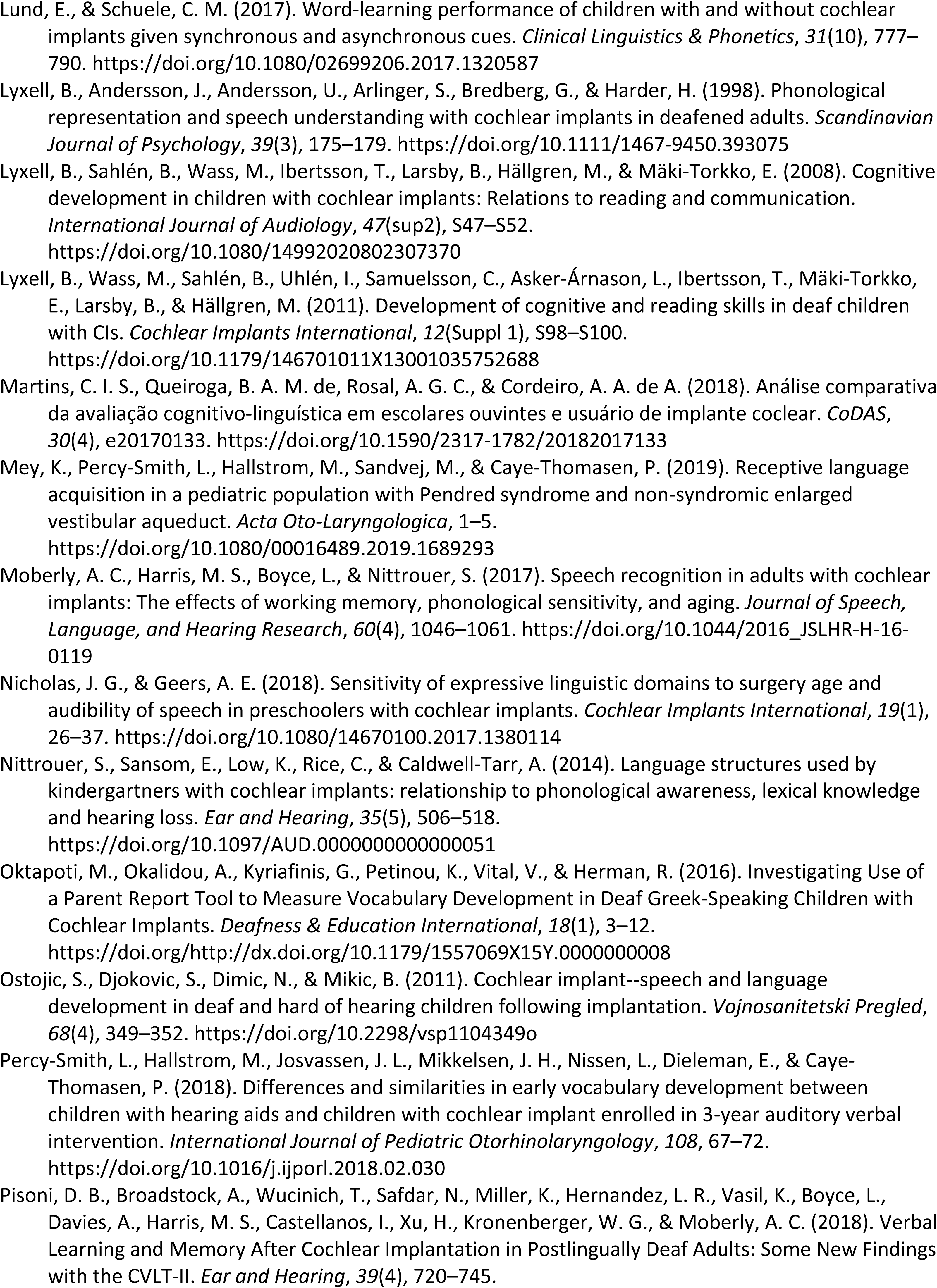
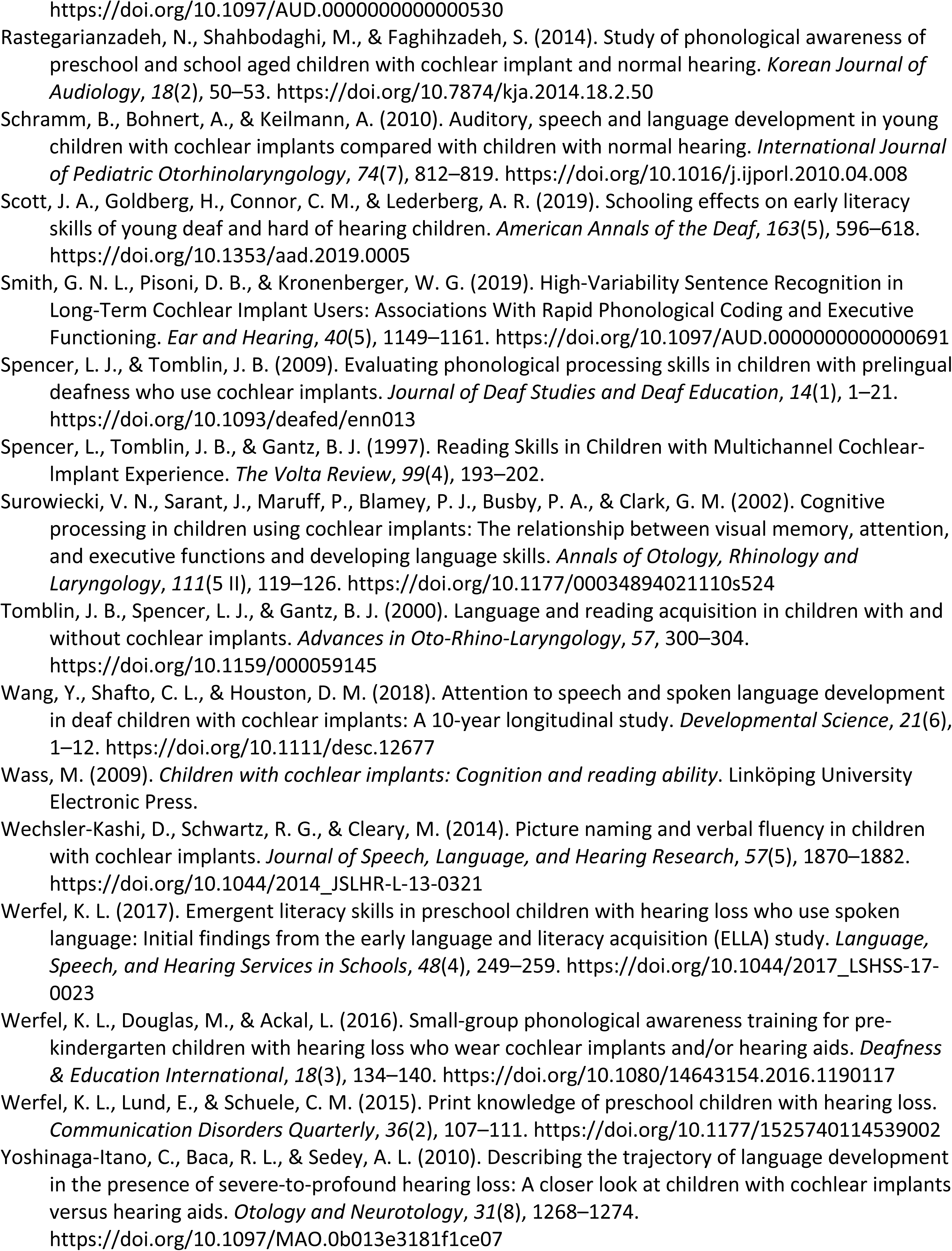
All 61 articles excluded after full-text screening in the current study

**Supplementary Table A.4.**
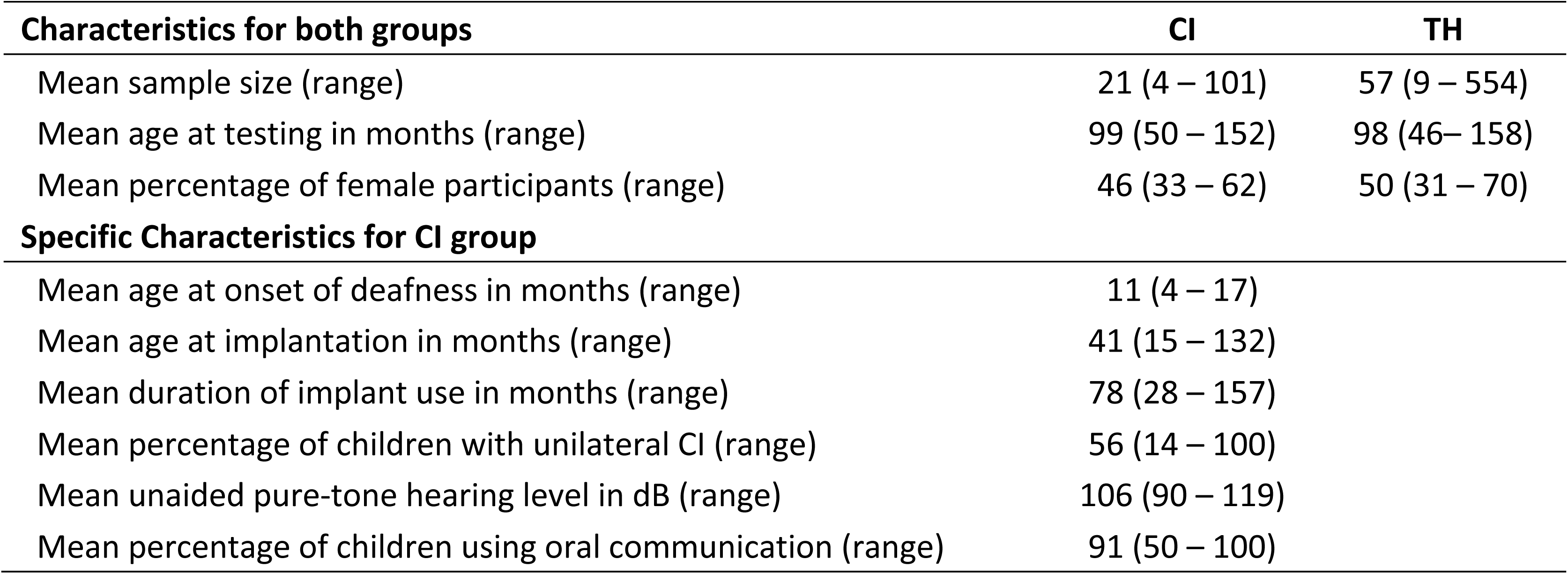
Demographic Information for CI versus TH

**Supplementary Table A.5.**
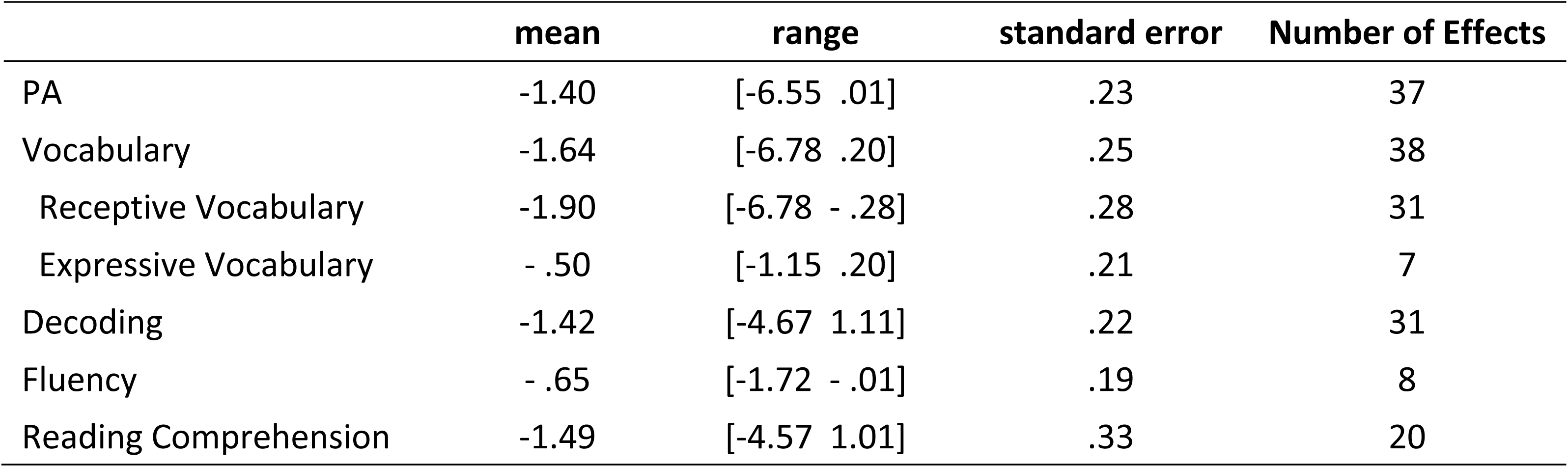
Descriptive Statistics of Hedges’ *g* for each construct for CI versus TH

**Supplementary Table A.6.** Demographic Information for CI versus HA

| **Characteristics for both groups** | **CI** | **HA** |
| --- | --- | --- |
| Mean sample size (range) | 24 (7 – 47) | 23 (7 – 71) |
| Mean age at testing in months (range) | 113 (62 – 165) | 115 (62 – 168) |
| Mean percentage of female participants (range) | 43 (33 – 61) | 54 (33 – 83) |
| **HL Characteristics for both groups** |  |  |
| Mean age at onset of deafness/HL in months (range) | 12 (6 – 19) | 14 (8 – 35) |
| Mean age at implantation/fitting in months (range) | 53 (20 – 129) | 28 (10 – 59) |
| Mean duration of device use in months (range) | 96 (37– 157) | 83 (83 – 83) |
| Mean percentage of children with unilateral CI (range)* | 56 (14 – 100) |  |
| Mean unaided pure-tone hearing level in dB (range) | 103 (93 – 117) | 88 (63 – 107) |
| Mean percentage of children using oral communication (range) | 79 (38 – 100) | 75 (33 – 100) |

\* many studies did not report bilateral HAs versus a unilateral HA. Thus, only the CI sample was reported.

## Supplementary Figure Legends

**Supplementary Figure A.1.**
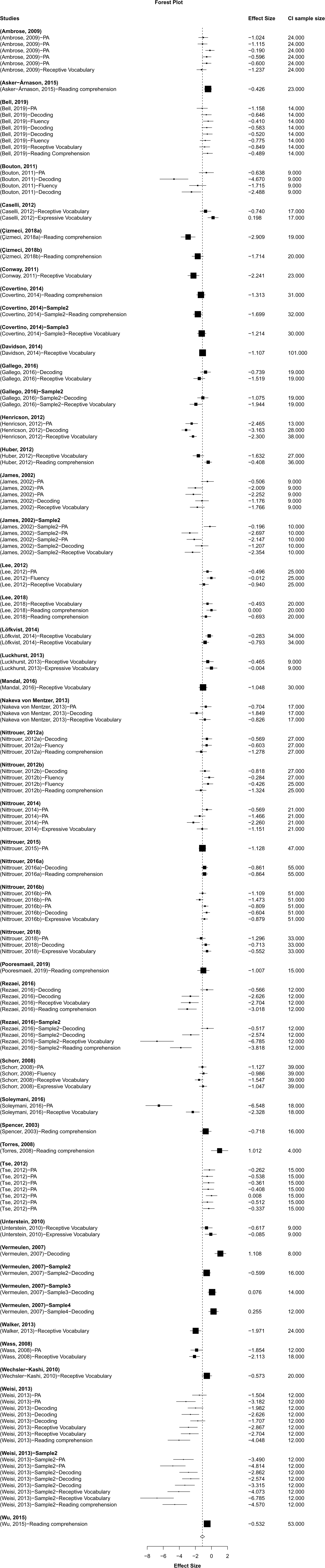
Forest Plot for CI versus TH

**Supplementary Figure A.2.**
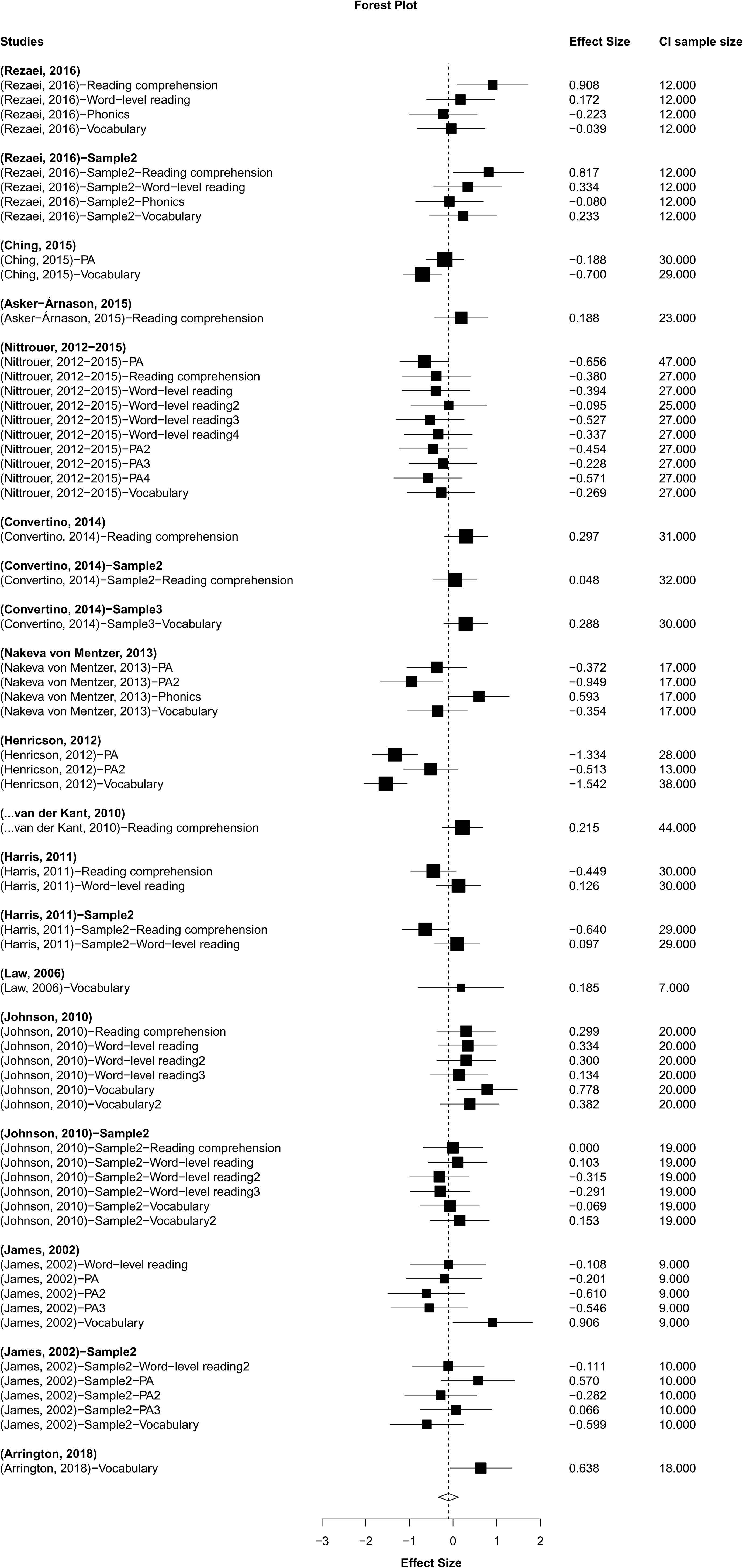
Forest Plot for CI versus HA

**Supplementary Figure A.3.**
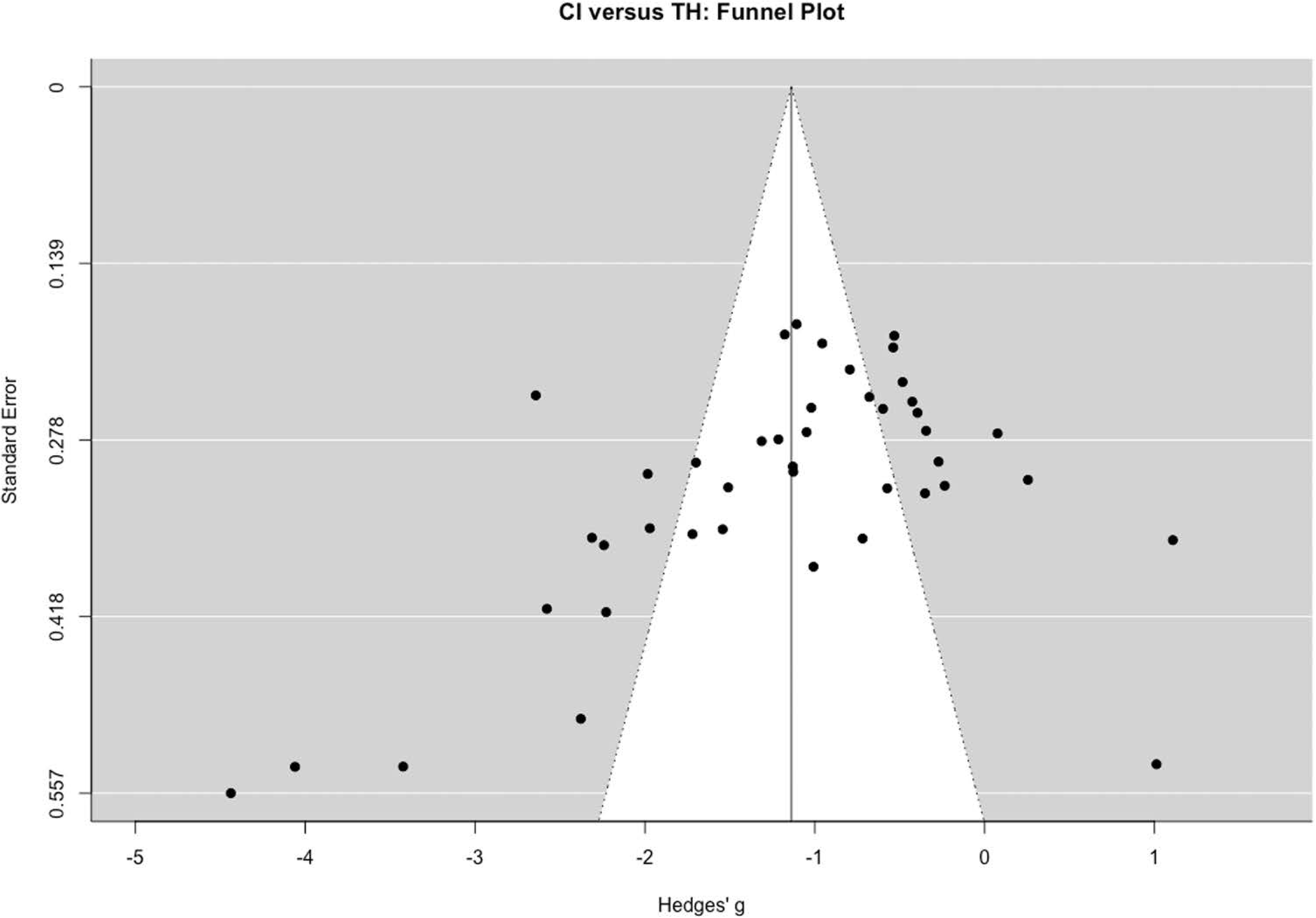
Funnel Plot for CI versus TH

**Supplementary Figure A.4.**
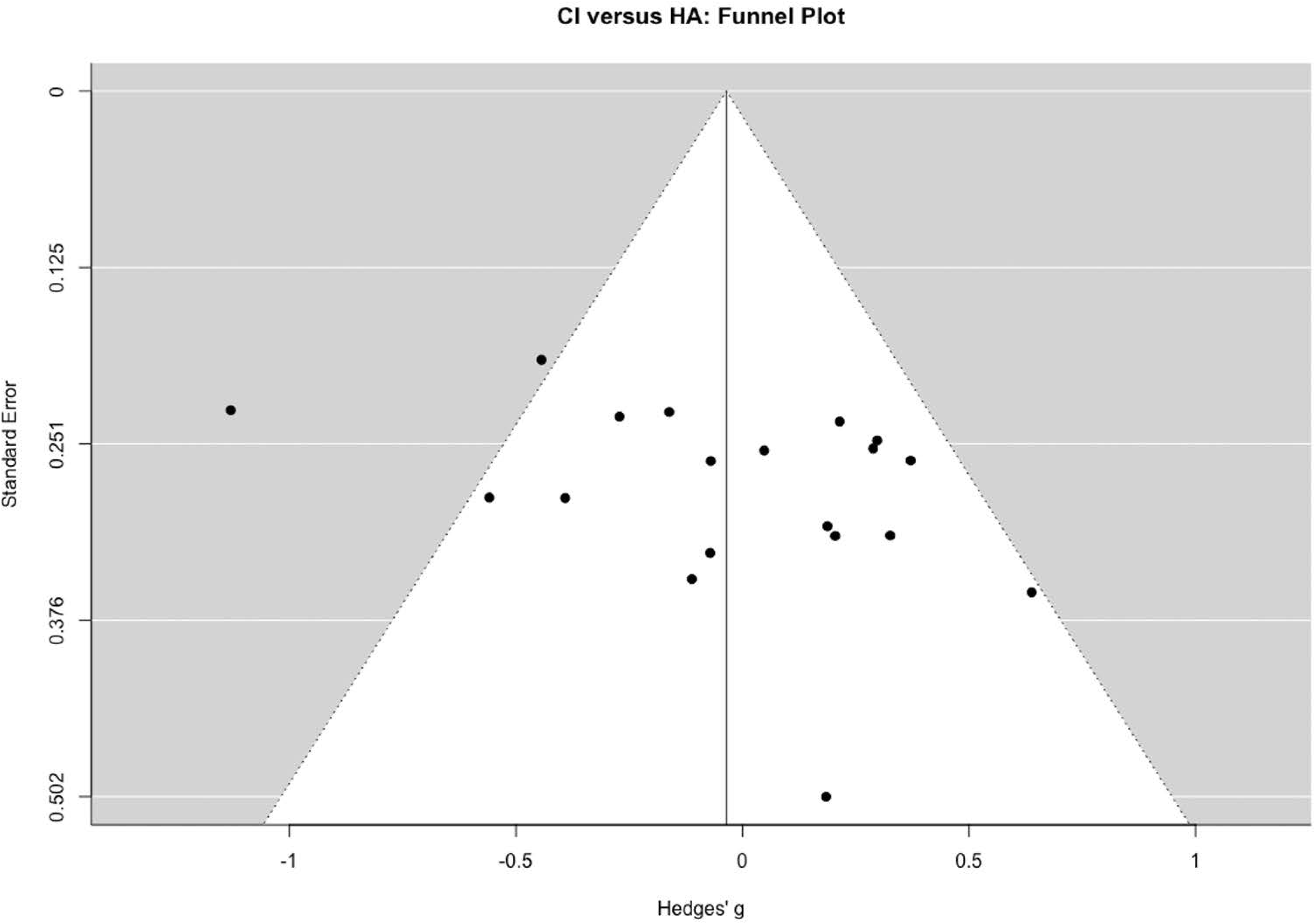
Funnel Plot for CI versus HA

